# Psychosis Prognosis Predictor: A Continuous and Uncertainty-Aware Prediction of Treatment Outcome in First-Episode Psychosis

**DOI:** 10.1101/2023.07.05.23292252

**Authors:** Daniël P.J. van Opstal, Seyed Mostafa Kia, Lea Jakob, Metten Somers, Iris E. C. Sommer, Inge Winter-van Rossum, René S. Kahn, Wiepke Cahn, Hugo G. Schnack

## Abstract

**Importance:** Presently, clinicians face challenges in accurately predicting the prognosis of patients with psychosis. Although machine learning models have shown promising potential in individual-level outcome prediction, their practical implementation as tools for real-world clinical practice has been hindered by several limitations. These limitations include difficulties in predicting multiple clinical outcomes, effectively capturing the evolving status of patients over time, and establishing trust in machine predictions. Addressing these shortcomings is crucial for responsibly leveraging machine learning in clinical decision-making for psychosis prognosis.

**Objective:** We propose and evaluate a multi-task recurrent neural network architecture specifically designed to overcome these challenges, enabling trustworthy prediction of psychosis prognosis. By developing a model that can effectively handle multiple clinical outcomes, capture the dynamic nature of patients’ status over time, and instill confidence in its predictions, we aim to provide clinicians with a robust tool for making informed and responsible decisions in the treatment of psychosis.

**Design, setting, and participants:** The study sample comprised 446 individuals, aged between 18 and 40 years, diagnosed with first-episode psychosis and participating in the OPTiMiSE study. To predict the likelihood of remission, we selected a range of multimodal baseline variables. These variables included PANSS scores (Positive and Negative Syndrome Scale), diagnostic subclass, duration of untreated psychosis, age at onset, sex, as well as physical health and lifestyle variables. The treatment outcome measures assessed at both week 4 and week 10 encompassed symptomatic remission, clinical global remission, and functional remission.

**Main outcome and measures:** we devised a recurrent neural network architecture incorporating long short-term memory (LSTM) units to facilitate outcome prediction by leveraging multimodal baseline variables and clinical data collected at multiple time points. To account for model uncertainty, we employed a novel fuzzy logic approach to integrate the level of uncertainty into individual predictions. To assess the predictive performance of our model, we employed rigorous evaluation techniques, including repeated 10-fold cross-validation and leave-one-site-out cross-validation.

**Results:** When solely utilizing pre-treatment patient status to forecast various outcomes in 4 weeks after starting the treatment, the leave-one-site-out validation process yielded area under the receiver operating characteristic curve (AUC) values ranging from 0.62 to 0.67. Notably, the predictive performance witnessed ∼0.04 improvement upon the inclusion of 1-week follow-up patient status, resulting in AUC values ranging from 0.66 to 0.72. Regarding the prediction of outcomes 10 weeks after the start of treatment, the models constructed solely with pre-treatment patient status achieved AUC values between 0.56 and 0.64. However, by incorporating follow-up patient status (specifically after 1, 4, and 6 weeks), the performance of the models was enhanced, resulting in AUC values of 0.72 to 0.74. Furthermore, after incorporating prediction uncertainties and stratifying the model decisions based on model confidence, we could achieve accuracies above 0.8 for ∼50% of patients in five out of six clinical scenarios.

**Conclusions and Relevance:** Our approach involved constructing prediction models utilizing a flexible neural network architecture tailored to clinical scenarios derived from a time series dataset. One crucial aspect we incorporated was the consideration of uncertainty in individual predictions, which enhances the reliability of decision-making based on the model’s output. Through our study, we provided compelling evidence showcasing the significance of leveraging time series data for achieving more accurate treatment outcome prediction in the field of psychiatry. Therefore, we advocate for the inclusion of time series data in future individualized prediction modeling endeavors within psychiatric research, as it holds substantial promise in improving prognostic accuracy and informing personalized treatment approaches.

## Introduction

There is an abundance of research into predictors of outcome in psychosis, but until now, clinicians are unable to reliably predict the disease course nor the success rate of (pharmacological) treatment intervention(s) of an individual patient. A possible way forward is the use of machine learning techniques^1, 2^. In psychiatry research, machine learning techniques are increasingly being used, particularly in psychotic disorders^3^. To the best of our knowledge, ten studies^4–13^ examined illness progress in existing psychotic disorders, each predicting different outcomes. Of these, only one study^4^ aimed to predict treatment response. In an open-label randomized clinical trial of five broadly used antipsychotics (*N* = 334), clinical and sociodemographic variables were used as inputs to a support vector machine to predict the level of functioning at four and 52 weeks after the start of antipsychotic treatment in patients with first-episode psychosis with an accuracy of 71– 72%.

The aforementioned psychosis prognosis prediction studies have noteworthy limitations that hinder their practical use as prediction tools in day-to-day clinical practice. Firstly, the importance of different outcomes may vary for individual patients, and therefore, clinicians and patients should have the ability to choose which outcomes are relevant for prediction. However, most existing prediction models in psychosis research are single-task models, focusing on predicting only one outcome measure. To overcome this limitation, we employ a multi-task learning^14^ design in our study, training a model to predict multiple outcome measures simultaneously, thus providing a more personalized and comprehensive prediction tool.

Secondly, in clinical practice, it is crucial to have an up-to-date prediction tool that can adapt to changes in a patient’s mental state and incorporate additional information obtained during each visit. Traditional machine learning methods used in psychosis prognosis prediction lack the ability to accommodate the dynamic nature of patients’ status. To address this limitation, we propose employing a machine learning approach capable of making predictions based on multiple assessments over time. One such approach is Long Short-Term Memory (LSTM), a type of recurrent neural network that has been successfully used in various healthcare domains^15^.

Thirdly, as machine learning models can be uncertain about their prediction (like human beings), the clinician needs to be informed about the uncertainty involved in model predictions. Knowing that the model is (very) sure about a certain prediction, the clinician may confidently integrate the prediction with their own judgment. On the other hand, when the tool is unsure about a certain prediction, the clinician may opt not to use the prediction as a guide to treat the patient. To date, most models do not provide the uncertainty in estimated model parameters (i.e., the epistemic uncertainty^16^), so it is unclear how far we can trust their predictions. Therefore it is desirable to integrate the model uncertainty in the predictions to facilitate the clinical usage of the model and to allow for more trustworthy decision-making^17, 18^. Such an improvement will reduce the risk of making wrong decisions, for example, by tapering or switching antipsychotic medication too early or unnecessarily late.

In our study, we present a machine learning framework that predicts multiple outcomes based on longitudinal patient data while integrating prediction uncertainty to facilitate more reliable clinical decision-making. This prediction model was trained using data from an international multicenter prospective clinical research trial. By addressing these limitations, we aim to enhance the applicability and trustworthiness of prediction models in guiding clinical practice and optimizing treatment strategies for patients with psychosis.

## Methods

### The dataset

Treatment of a first-episode patient is a sequence of (re)evaluation of the patient’s status and effects of treatment thus far and decisions about (changing) treatment. At each time point, the psychiatrist integrates newly available data with information gathered in the past. For it to be useful in this clinical practice, a machine-learning prediction tool must do the same. In this study, the functioning of the prediction model is evaluated in the OPTiMiSE study^19^, a large, international, multicenter antipsychotic three-phase switching study in which patients with first-episode psychosis were examined at multiple visits and treated with antipsychotic medication that could be changed based on the patient’s response. We used data from patients in phase one and phase two. In the first phase, patients (N=446/371 started/completed) were treated with amisulpride (up to 800 mg/day) for four weeks. Patients who then met the criteria for symptomatic remission did not continue to the next phase. Patients not in remission went on to phase two (N=93/72 started/completed) and either continued using amisulpride or switched to olanzapine (≤20 mg/day) for six weeks.

Our primary outcome measure for prediction was symptomatic remission. Secondary outcome measures were clinical global remission and functional remission. Symptomatic remission was defined the same way as in the OPTiMiSE study, according to the consensus criteria of Andreasen et al.^20^ based on the Positive And Negative Syndrome Scale (PANSS)^21^, albeit without the minimum duration of six months. For global illness, we used the Clinical Global Impression (CGI) scale^22^. We considered a CGI score of 4 or lower as clinical global remission. For the functional outcome, we used the Personal and Social Performance (PSP) scale. We considered a global PSP score of 71 points or higher as functional remission, following Morosini’s definition where a global PSP score from 71 to 100 points refers only to mild difficulties^23^ (for an overview of all included predictor variables, see Table 1).

**Table 1:**
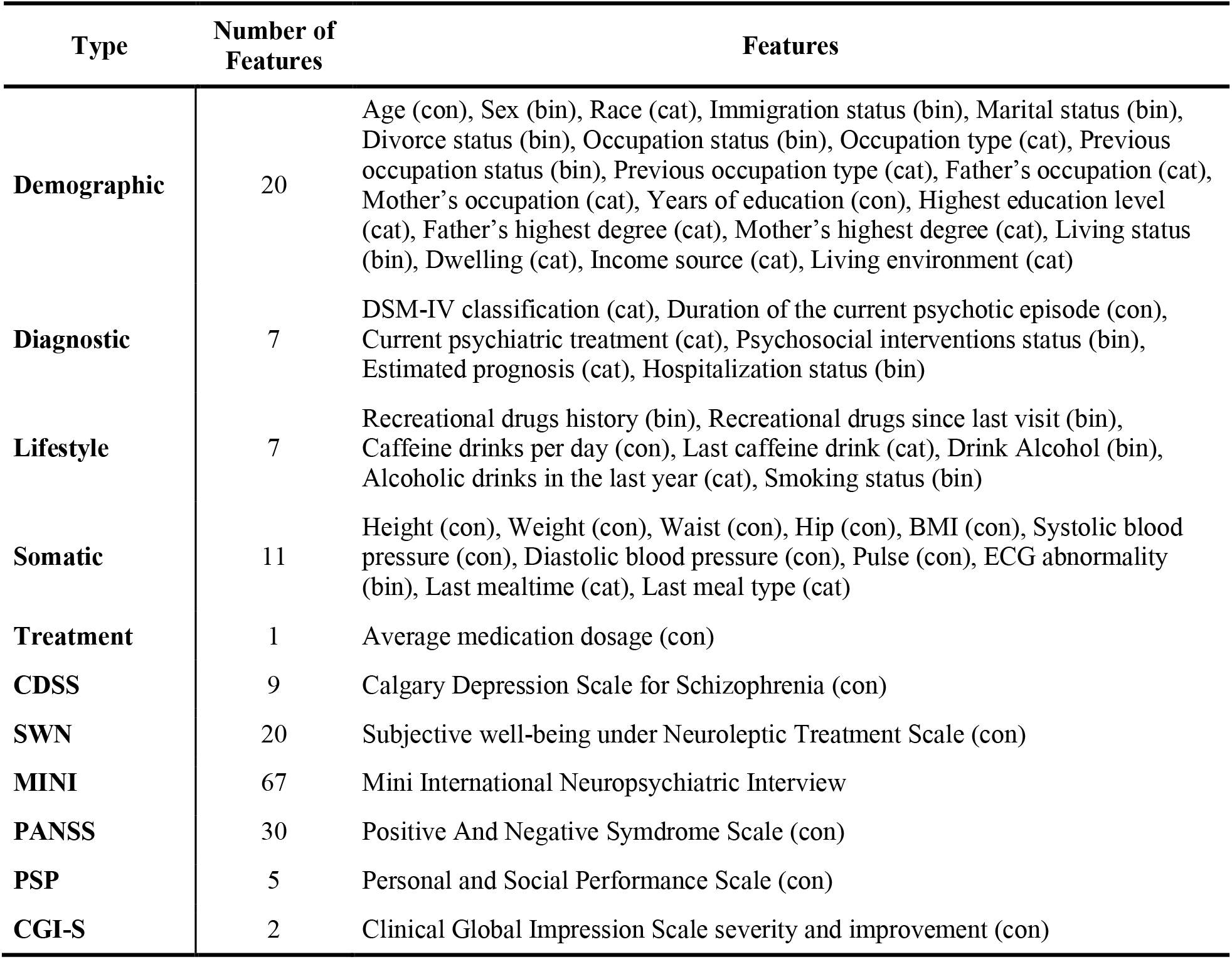
The type, number, and list of features from OPTiMiSE study that are used as predictors in our model. con: continuous measure, bin: binary measure, cat: categorical measure.

| Type | Number of Features | Features |
| --- | --- | --- |
| **Demographic** | 20 | Age (con), Sex (bin), Race (cat), Immigration status (bin), Marital status (bin), Divorce status (bin), Occupation status (bin), Occupation type (cat), Previous occupation status (bin), Previous occupation type (cat), Father's occupation (cat), Mother's occupation (cat), Years of education (con), Highest education level (cat), Father's highest degree (cat), Mother's highest degree (cat), Living status (bin), Dwelling (cat), Income source (cat), Living environment (cat) |
| **Diagnostic** | 7 | DSM-IV classification (cat), Duration of the current psychotic episode (con), Current psychiatric treatment (cat), Psychosocial interventions status (bin), Estimated prognosis (cat), Hospitalization status (bin) |
| **Lifestyle** | 7 | Recreational drugs history (bin), Recreational drugs since last visit (bin), Caffeine drinks per day (con), Last caffeine drink (cat), Drink Alcohol (bin), Alcoholic drinks in the last year (cat), Smoking status (bin) |
| **Somatic** | 11 | Height (con), Weight (con), Waist (con), Hip (con), BMI (con), Systolic blood pressure (con), Diastolic blood pressure (con), Pulse (con), ECG abnormality (bin), Last mealtime (cat), Last meal type (cat) |
| **Treatment** | 1 | Average medication dosage (con) |
| **CDSS** | 9 | Calgary Depression Scale for Schizophrenia (con) |
| **SWN** | 20 | Subjective well-being under Neuroleptic Treatment Scale (con) |
| **MINI** | 67 | Mini International Neuropsychiatric Interview |
| **PANSS** | 30 | Positive And Negative Syndrome Scale (con) |
| **PSP** | 5 | Personal and Social Performance Scale (con) |
| **CGI-S** | 2 | Clinical Global Impression Scale severity and improvement (con) |

### The Psychosis Prognosis Predictor

In clinical practice, patients undergo multiple assessments throughout the treatment phase to monitor their progress. Similarly, in the OPTiMiSE study, patients were assessed at various time points during different phases of the study. These assessments included baseline (week 0, W_0_), the end of phase one (week four, W_4_), and the end of phase two (week ten, W_10_), as well as additional assessments at weeks one, two, six, and eight (W_1_, W_2_, W_6_, and W_8_, respectively). These frequent assessments allow for a comprehensive evaluation of a patient’s status and enable the tracking of changes over time. By incorporating data from these multiple time points, our study aims to capture the dynamic nature of the disease and improve the accuracy of psychosis prognosis prediction.

To address the objective, we introduce a multi-modal, time-aware, and multi-task recurrent neural network architecture designed specifically for psychosis prognosis prediction. This architecture is capable of handling multi-modal data from various sources, capturing the dynamic nature of the data as it evolves over time, and simultaneously predicting multiple outcome measures. The proposed architecture, depicted in Figure 1, comprises four conceptual modules that work synergistically to achieve accurate predictions (for a detailed description, see eMethods in the supplementary materials):

1. **Static module:** which is responsible for handling the static input features that remain constant over time. This module preprocesses the static features through a specific procedure tailored to different types of features. For continuous features, the missing data is imputed using the median value of that feature. Subsequently, a robust min-max scaler is applied to rescale the features within the range of [0,1]. To ensure the scaler is robust to outliers, the min and max values are determined based on the median of the bottom and top 10 percent of the feature values, respectively. Binary and categorical features undergo a similar preprocessing procedure. Missing values in binary and categorical features are replaced with the most frequent values observed in the dataset. Categorical features are then subjected to one-hot encoding to represent the different categories, while binary variables are encoded using a simple –1/1 scheme. By employing these preprocessing steps within the static module, the input static features are effectively prepared for integration with the dynamic components of the architecture.
2. **Dynamic module:** which is responsible for processing the input data that changes over time, known as the dynamic features. In this study, the dynamic measures consist of PANSS (30 measures), PSP (5 measures), and CGI (2 measures) assessments. The dynamic module consists of modality-specific long short-term memory (LSTM) units, a recurrent neural network architecture^24^, that is well suited for making predictions on time series data^25, 26^. The number of neurons in each LSTM unit is determined by multiplying the number of measures in each assessment by 2; thus, 60 neurons for PANSS, 10 neurons for PSP, and 4 neurons for CGI. The LSTM units serve as the representation learning layer within the dynamic module. They receive the dynamic features starting from the baseline (W_0_) up to a user-defined endpoint *t*. The LSTM units process the dynamic features and transform them into time-varying middle representations of dynamic data, denoted as Z_1_, Z_2_, and Z_3_ for PANSS, PSP, and CGI measures, respectively. After the dynamic features are transformed into these learned representations, they are merged with the static features in the fusion layer. This fusion step combines static and dynamic information, resulting in a comprehensive set of features. The fused features are then forwarded to the subsequent layers in the regression and classification modules for further analysis and prediction.
3. **Regression module:** which receives the outputs of the static and dynamic modules to predict the dynamic data at the next time point *t*+1. It takes the merged static features and learned dynamic representations as inputs and employs modality-specific two-layer dense layers with ReLU (rectified linear unit) transfer function for this mapping. The first layer in the regression module is called the interaction layer. It consists of time-distributed dense layers that have 30, 10, and 4 neurons respectively for PANSS, PSP, and CGI measures. The purpose of this layer is to capture the potential interactions between different static and dynamic data modalities. The number of neurons in this layer is determined based on the number of features in each modality, allowing for the computation of residuals. To compute the residuals, the outputs of the interaction layers are subtracted from the actual PANSS, PSP, and CGI values in the inputs. The second layer in the regression module is the output layer, which predicts the dynamic features at the next time point. By being able to predict the dynamic features of the subsequent time point, the model gains the capability to predict the future indefinitely. This is achieved by concatenating the predicted outputs with the dynamic inputs and feeding them back into the network for predicting the measures at *t+2* (the thick yellow arrow in Fig. 1). This recursive procedure can be utilized to predict the model’s output at time *t+k,* where *k* represents an arbitrary time point in the future, given the data from time 0 to time *t*.
4. **Classification module:** this receives the same inputs as the regression module and predicts the probability of target classes (not-remitted or remitted) at time *t+*1 for three outcome measures namely symptomatic remission (SR), clinical global remission (CR), and functional remission (FR). In the interaction layer of the classification module, task-specific and time-distributed dense layers with 5 neurons and ReLU transfer functions are utilized to capture the interactions between features. The number of neurons in this layer is predetermined prior to experiments to ensure a fair evaluation and does not undergo further optimization. This layer aims to exploit the potential interactions between different features. The output layer consists of *softmax* layers with two neurons for each task. These neurons predict the probability of the target classes for each outcome measure, distinguishing between the remitted and not-remitted states. By employing a multi-task architecture in the classification module, the network can leverage the covariance structure that may exist among the outcome variables. To enhance the reliability of the predicted probabilities in the classification module, isotonic regression is applied to the training data. This additional calibration process allows for the direct interpretation of the predicted probabilities as a confidence level for each class in the outcomes and improves the reliability of the confidence estimates.

**Figure 1.**
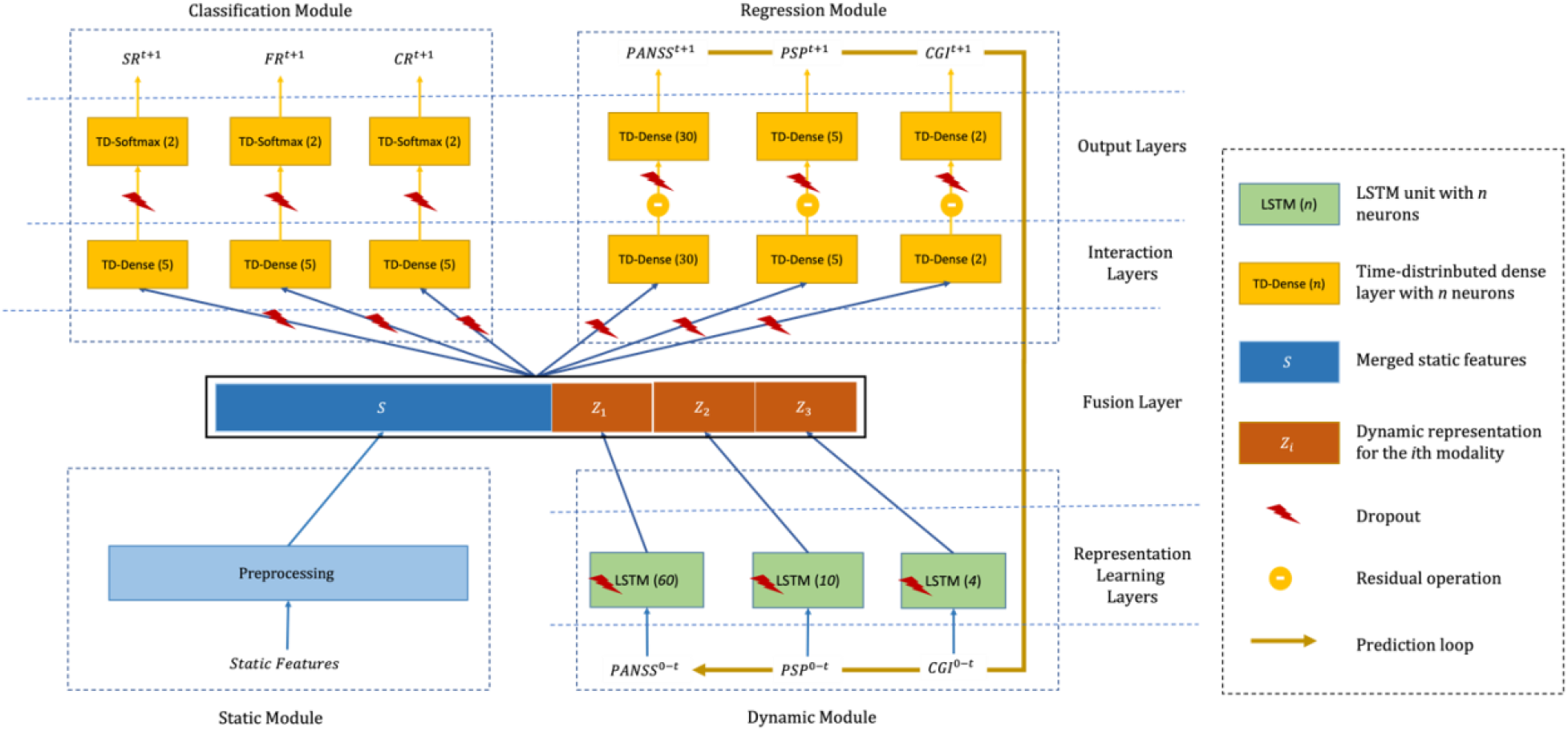
**The psychosis prognosis predictor architecture consists of four layers that are organized into four conceptual modules**. The layers include 1) the representation learning layer that learns a middle representation for dynamic features; 2) the fusion layer that merges the preprocessed static features with dynamic middle representations; 3) the interaction layer that seeks to benefit from interaction between static features and dynamic features from different modalities; 4) the output layer that predicts the outputs at the next time step. The modules include 1) the static module for preprocessing and merging the static features; 2) the dynamic module that includes LSTM units for learning middle representation for dynamic features from time *0* to time *t*; 3) the regression module for predicting the dynamic measures at the next time step (*t+1*), and 4) classification module for predicting the outcomes (*SR*: symptomatic remission, *FR*: functional remission, *CR*: clinical remission) at the next time step. The prediction loop from the output of the regression module to the inputs of the dynamic module (the thick yellow arrow) enables the network to predict the outcomes at an arbitrary future point.

### Pretraining, data augmentation, and training

The proposed architecture has 39,870 parameters, while the number of available samples in the OPTiMiSE dataset is ∼400 patients. This makes the optimization process challenging. To address this problem, we used pretraining and data augmentation techniques. In this direction, we first simulated the data for 10000 patients by randomly drawing from the range between the minimum and maximum values for each feature. Then, we pre-trained the network using simulated synthetic data. For data augmentation, we augmented the samples in the training set using a variable-length sliding window. Figure 2 illustrates the data augmentation procedure in which 10 samples with time lengths of 2, 3, 4, and 5 are derived from one sample with a time length of 5.

**Figure 2.**
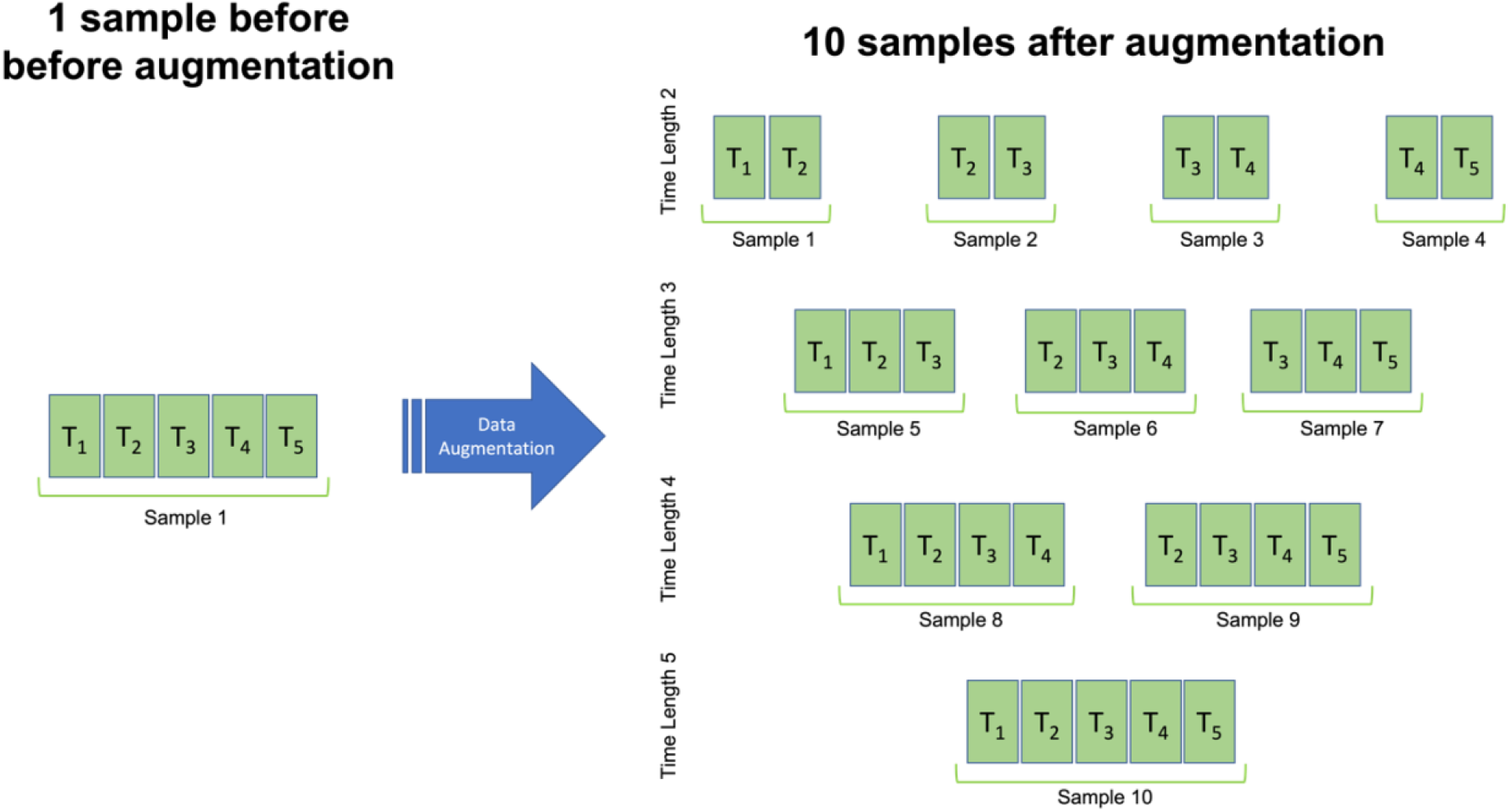
**The data augmentation process**. A set of ten samples with time-length 2 to 5 are generated for a sample with the length of five timepoints.

Furthermore, we used dropout, with a two-fold advantage: during the training, it prevents the network from overfitting^30^; while in the prediction phase, it enables estimating the uncertainty in the predictions^31^. The estimated uncertainties are used in the proposed decision-making module (see section **From predictions to uncertainty-aware clinical decisions**) to translate the model predictions of outcomes into risk-aware clinical decisions.

A multi-objective loss function is optimized during the training procedure in which weighted mean squared and categorical cross-entropy loss functions are respectively used for regression and classification tasks. We used 1 and 5 as weights respectively for regression and classification loss functions. An Adam optimizer with an initial learning rate of *3* × *10*^−*4*^ was employed during the optimization procedure. The exponential learning rate decay with a decay rate of 0.9 and decay steps of 10000 is used. In the pretraining phase, we trained the network for two epochs with a mini-batch size of 25 samples. In the training phase, the network is trained on augmented data for 50 epochs with a mini-batch size of 2 samples.

### From predictions to uncertainty-aware clinical decisions

In general, the probabilities predicted by a classifier are used as outcomes for clinical decision-making, by discretizing the probabilities into classes of decisions by imposing a hard threshold (e.g., 0.5 for two classes). However, a classifier, like a human being, can sometimes be unsure about its predictions. How sure a model is about its predictions can be quantified by estimating the epistemic uncertainty^16, 27^, i.e., the uncertainty in the model parameters. Now, the challenge is to combine the predicted probabilities and their estimated uncertainties into final clinical decisions.

In this paper, we use a fuzzy logic^28^ approach for translating the predictions of the model into uncertainty-aware clinical decisions. Fuzzy logic provides a mathematical framework for representing vague and imprecise information. We employ Mamdani’s rule-based fuzzy inference procedure^29^ in four steps:

1. **Uncertainty interval estimation:** 100 repetitions of the Monte-Carlo dropout technique ^31^ are used to estimate the uncertainties in predictions of the classification module. The median value across these 100 predictions is used as the estimated probability of remission (*p*). Then, the robust minimum and maximum (median of 10% lowest/highest) predicted probabilities are calculated to estimate the worst-case and the best-case probability of remission (*p_w_* and *p_b_*), respectively. In fact, *p_b_* minus *p_w_* represents the uncertainty interval for a certain prediction.
2. **Fuzzification of inputs and outputs:** Fuzzification is the process of translating numerical values into linguistic variables. This is performed via fuzzy membership functions that assign a degree of membership (between 0 and 1) to each numerical value. In our specific application, we define 5 Gaussian membership functions (Figure 3a) for the predicted probabilities (*p*, *p_w_*, and *p_b_*), including *G*(*0*, σ) for ‘very low’, *G*(*0*.*25*, σ/*2*) for ‘low’, *G*(*0*.*5*, σ) for ‘medium’, *G*(*0*.*75*, σ/*2*) for ‘high’, and *G*(*1*, σ) for ‘very high’. Furthermore, 7 Gaussian membership functions (Figure 3b) are used to categorize the outcome decisions into 7 categories, including *G*(*0*, σ/*2*) for ‘definite no-remission (DN)’, *G*(*0*.*2*, σ/*2*) for ‘probable no-remission (PN)’, *G*(*0*.*4*, σ/*2*) for ‘unsure no-remission (UN)’, *G*(*0*.*5*, σ/*4*) for ‘unsure (US)’, *G*(*0*.*6*, σ/*2*) for ‘unsure remission (UR)’, *G*(*0*.*8*, σ/*2*) for ‘probable remission (PR)’, and *G*(*1*, σ/*2*) for ‘definite remission (DR)’. In our experiments, since we have 7 decision categories, we heuristically set σ = *1*/*7* = *0*.*143*.
3. **Fuzzy inference:** where a set of seven if-then rules are used to translate the membership values for input probability of remissions (*p*, *p_w_*, and *p_b_*) into membership values for seven decision categories (DN, PN, UN, US, UR, PR, and DR). The logical *‘and’* and *‘or’* operations within the rules are respectively implemented using fuzzy *‘max’* and *‘min’* operators. Figure 4 depicts the fuzzy inference procedure using these seven if-then rules when *p*=0.90, *p_w_*=0.25, and *p_b_*=1.00. For example, the first rule says ‘if the probability of remission is very high and the worst-case probability of remission is high or very high, then the decision is definite remission’. The membership value of a very high probability of remission for *p*=0.90 is 0.78, and membership values of the high and very high worst-case probability of remission for *p_w_*=0.25 are approximately 0. Thus, the membership value for the definite remission decision is *min*(*0*.*78*, *max*(*0*,*0*)) = *0*. In this example, PR and UR decisions have the highest membership values with 0.21 and 0.78, respectively, and the memberships for the rest categories remain very close to zero. We use the maximum membership value across seven outcome decisions to specify the final decision. In this case, the final decision is unsure remission (UR) which is a reasonable decision given that the predicted probability of remission is high but the model is very uncertain about it.
4. **Aggregation and defuzzification:** to calculate the uncertainty-aware probability of remission 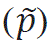 given the model uncertainty, we first aggregate the membership functions for seven clinical decisions using the fuzzy max aggregation that performs similarly to a union operation. Then in the defuzzification step, we compute the centroid of the aggregated mass and use its *x* coordinate as the modified probability of remission. In our example, the uncertainty-aware probability of remission is 0.65 which shows a significant discount compared to the initial probability of remission of 0.90 given its high uncertainty.

**Figure 3.**
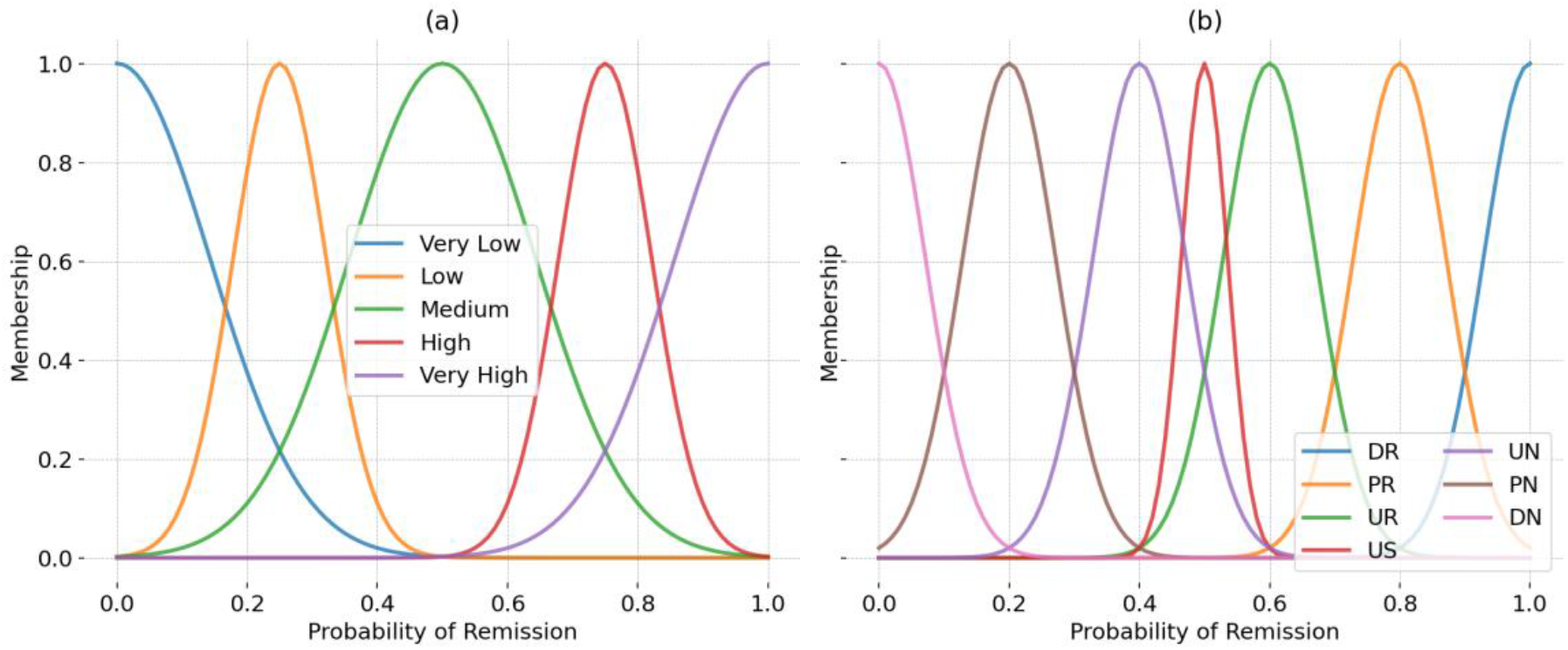
a) Five Gaussian membership functions for the probability of remission. These functions are used to map the values of the probability of remission (*p*), the worst-case probability of remission (*p_w_*), and the best-case probability of remission (*p_b_*) in the x-axis to a membership value (between 0 and 1) in the y-axis for ‘very low’, ‘low’, ‘medium’, ‘high’, and ‘very high’ categories; b) Gaussian membership functions for seven clinical decisions, ‘definite no-remission (DN)’, probable no-remission (PN)’, ‘unsure no-remission (UN)’, ‘unsure (US)’, ‘unsure remission (UR)’, ‘probable remission (PR)’, ‘definite remission (DR)’.

**Figure 4.**
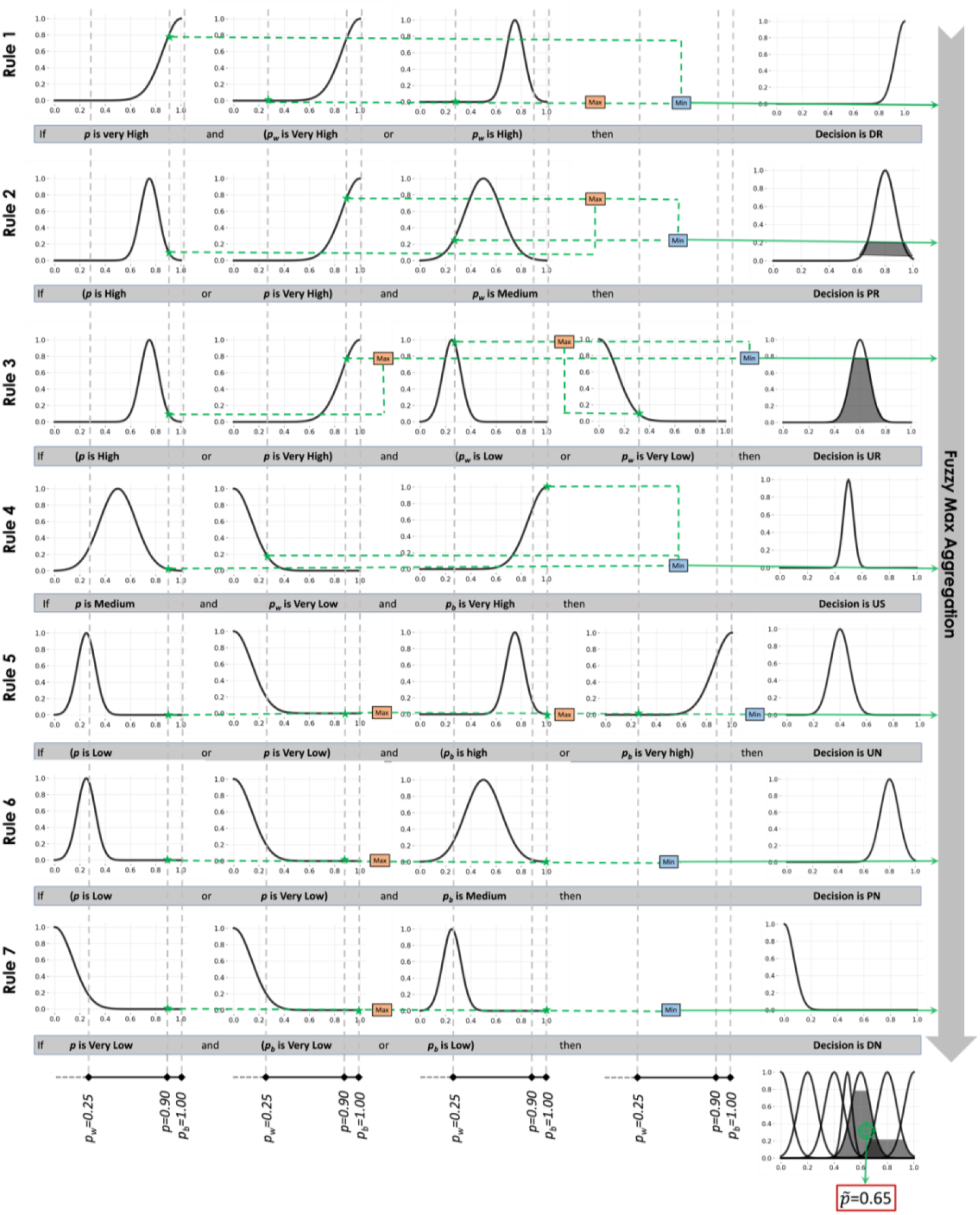
Seven rules in the proposed fuzzy inference system for translating the predicted probability of remission (*p*), the worst-case probability of remission (*p_w_*), and the best-case probability of remission (*p_b_*) into risk-aware clinical decisions. The green stars show the value of the corresponding membership function in each rule for an example prediction with *p*=0.9, *p_w_*=0.25, and *p_b_*=1.00. The orange and blue boxes represent the fuzzy max and min operations, respectively. The gray area in the last right column shows the mass under the membership function of each decision. These masses are combined using fuzzy max aggregation. The x-coordinate of the centroid of the aggregated mass represents the uncertainty-aware probability of remission 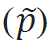 that aggregates the model uncertainty into the final prediction.

Figure 5 shows how the fuzzy logic framework modifies the predicted probability of remission based on the estimated model uncertainty. Figure 5b shows how the decision surface is divided between seven categories of decisions. These uncertainty-aware categorical decisions can play the role of meta-information aiding clinicians in more reliable AI-aided decision-making. For example, if a decision lies in one of the “unsure” categories, the clinicians can ignore the model prediction and rely on other sources of information (e.g., a second opinion from a colleague or gathering more information about the patient).

**Figure 5:**
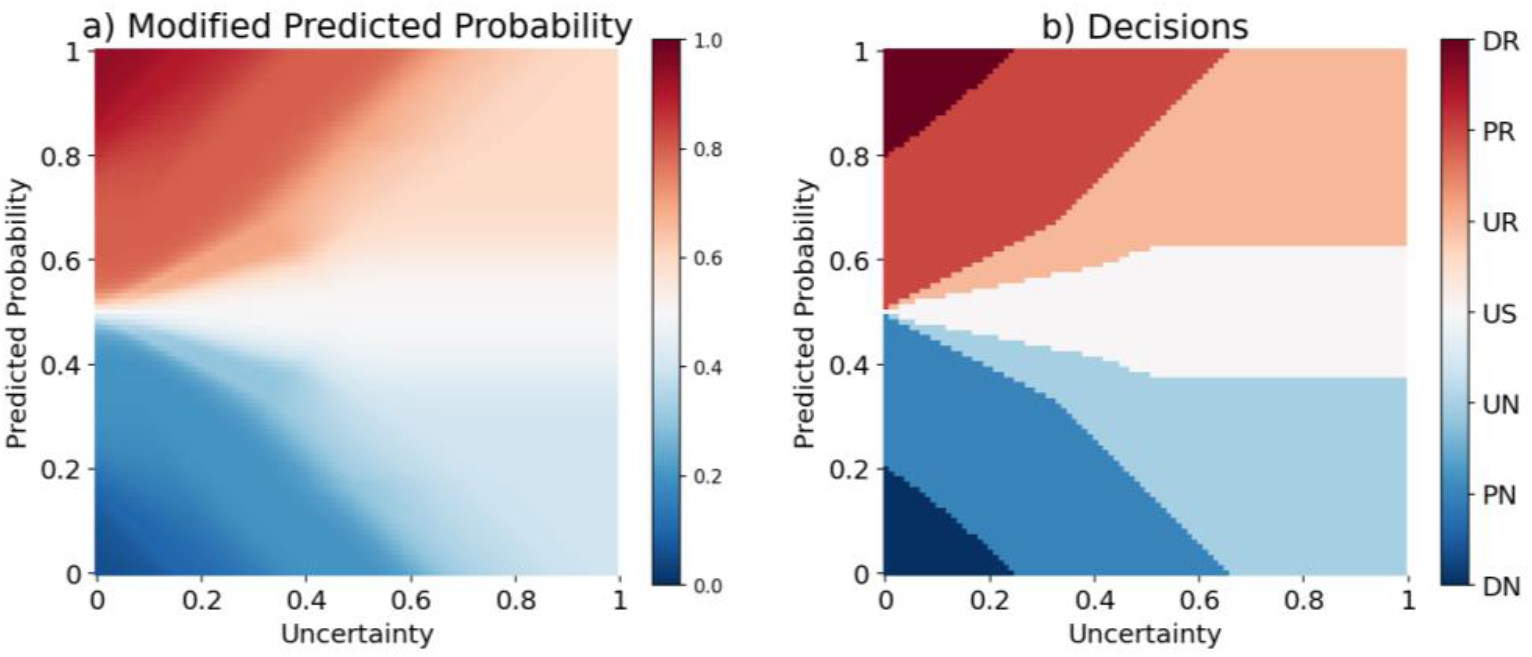
**a) the modified probability of symptomatic remission (color scale) after adjusting the predicted probability (y-axis) based on model uncertainty (x-axis)**. For example, *p* = 1.00 (i.e., the model predicts 100% remission for a certain patient) can be transferred to a value between 0.60 to 1.00 depending on the level of model uncertainty. This modification resulted in better-calibrated predictions of the probability of remission and is thus more suitable for clinical usage (see eResults). Calibration of the prediction models is a critical factor but often neglected^33^, especially in identifying the threshold of risks in clinical decision-making^34^. **b) the span of decision surfaces for seven categories of clinical decisions.** The fuzzy logic approach for decision-making enables the prediction model to also say “I do not know” when the decision lies in the unsure (US) category. This is a crucial feature for more safe applications of ML models in clinical settings^17^. Furthermore, psychiatrists can refrain from relying on model predictions when the decisions lie in the unsure remission (UR) or unsure no-remission (UN) to reduce the risk of wrong decisions.

### Model evaluation

In the study, the classification performance of the proposed architecture was evaluated using 20 repetitions of two cross-validation procedures: 10-fold cross-validation and one-site-out cross-validation. The repeated cross-validation procedures helped account for variations and ensure reliable estimates of the model’s performance. For each repetition of the cross-validation, evaluation metrics were calculated to measure the classification performance. The metrics used in this study include

1) Area Under the Receiver Operating Characteristic Curve (AUC): This metric quantifies the overall discriminative power of the model. It represents the ability of the model to distinguish between the positive and negative classes.
2) Balanced Accuracy (BAC): BAC takes into account both sensitivity and specificity and provides a balanced measure of the model’s accuracy. It is particularly useful when the dataset is imbalanced, meaning the number of samples in each class is significantly different.
3) Sensitivity: Sensitivity measures the proportion of true positive predictions out of all actual positive samples. It indicates the model’s ability to correctly identify positive cases.
4) Specificity: Specificity measures the proportion of true negative predictions out of all actual negative samples. It indicates the model’s ability to correctly identify negative cases.

### Experimental setup

We use the model to predict the outcomes at four weeks (W_4_) and ten weeks (W_10_) following the initiation of treatment (W_0_). In order to assess the impact of including patient status information obtained during the treatment phase on the accuracy of the predictions, we conducted a performance comparison of the predictor using different lengths of data points over time, ranging from W_1_ to W_6_ (as illustrated in Figure 6). This evaluation was carried out across six distinct clinical scenarios (S_1_-S_6_), allowing us to examine the predictive capabilities of the model under various conditions.

**Figure 6:**
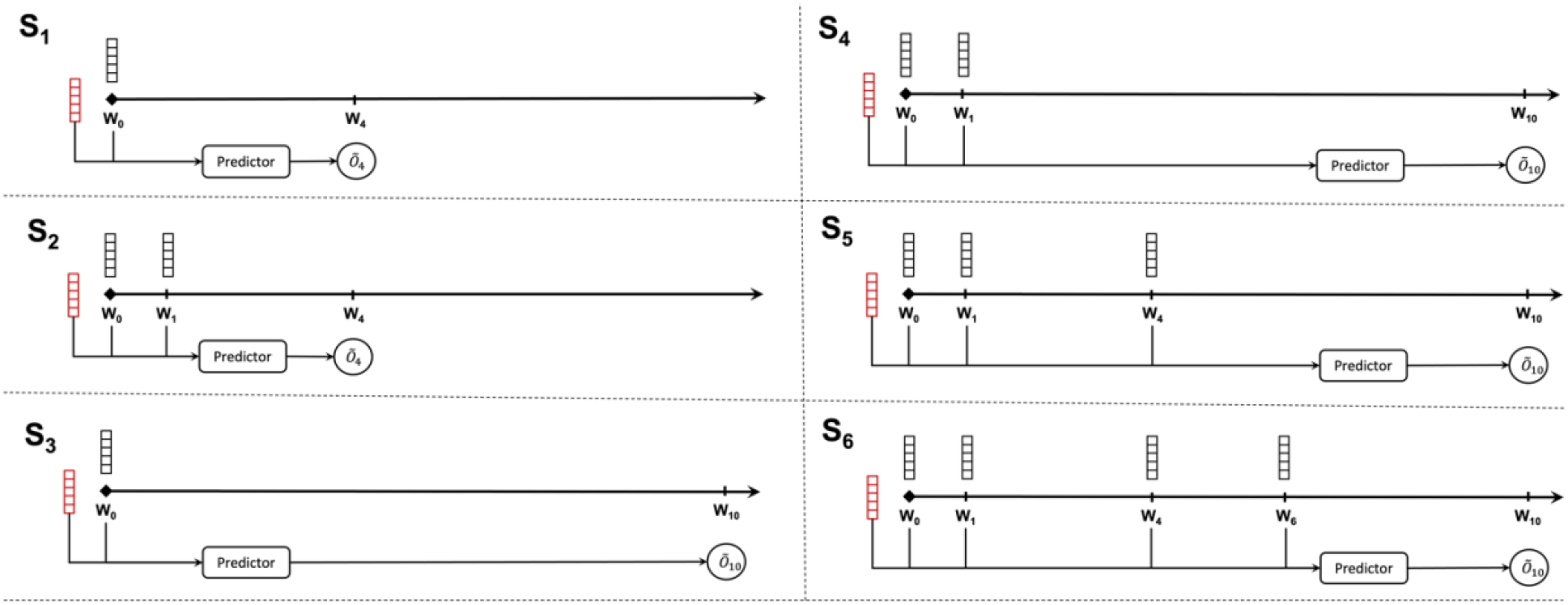
prognosis prediction models in six clinical scenarios. Static and dynamic features are represented as red and black blocks, respectively. **(S_1_)** Only information from W_0_ is used for predicting the outcome at W_4_. **(S_2_)** Prediction at W_4_ is made after a 1-week follow-up by adding the dynamic information from W_1_. **(S_3_)** Only information from W_0_ is used for predicting the outcome at W_10_. **(S_4_)** Prediction at W_10_ is made after a 1-week follow-up by adding the dynamic information from W_1_. **(S_5_)** All the information from the first phase of the study is used to predict the outcome at W_10_. **(S_6_)** The dynamic information from W_6_ is also used as input data to the model for prediction at W_10_.

## Results

### More data over time results in higher prediction accuracy

As summarized in Table 2 and Figure 7, using one-site-out cross-validation, AUC for the 4-week outcomes in S_1_ and S_2_ scenarios ranged from 0.66 for functional remission to 0.71 for symptomatic remission. For the 10-week outcomes, AUC ranged from 0.72 for functional remission to 0.74 for symptomatic remission (for balanced accuracy, sensitivity and specificity, see sTable 1-3 and sFigure 1-3 in the supplementary materials). Across all outcome measures, the AUC of the 4-week predictions improved by 0.04–0.05 when not only baseline data (W_0_) but also data after one week (W_1_) was used. For 10-week predictions, the use of all time series data improved AUC by 0.08–0.17, across all outcome measures.

**Figure 7.**
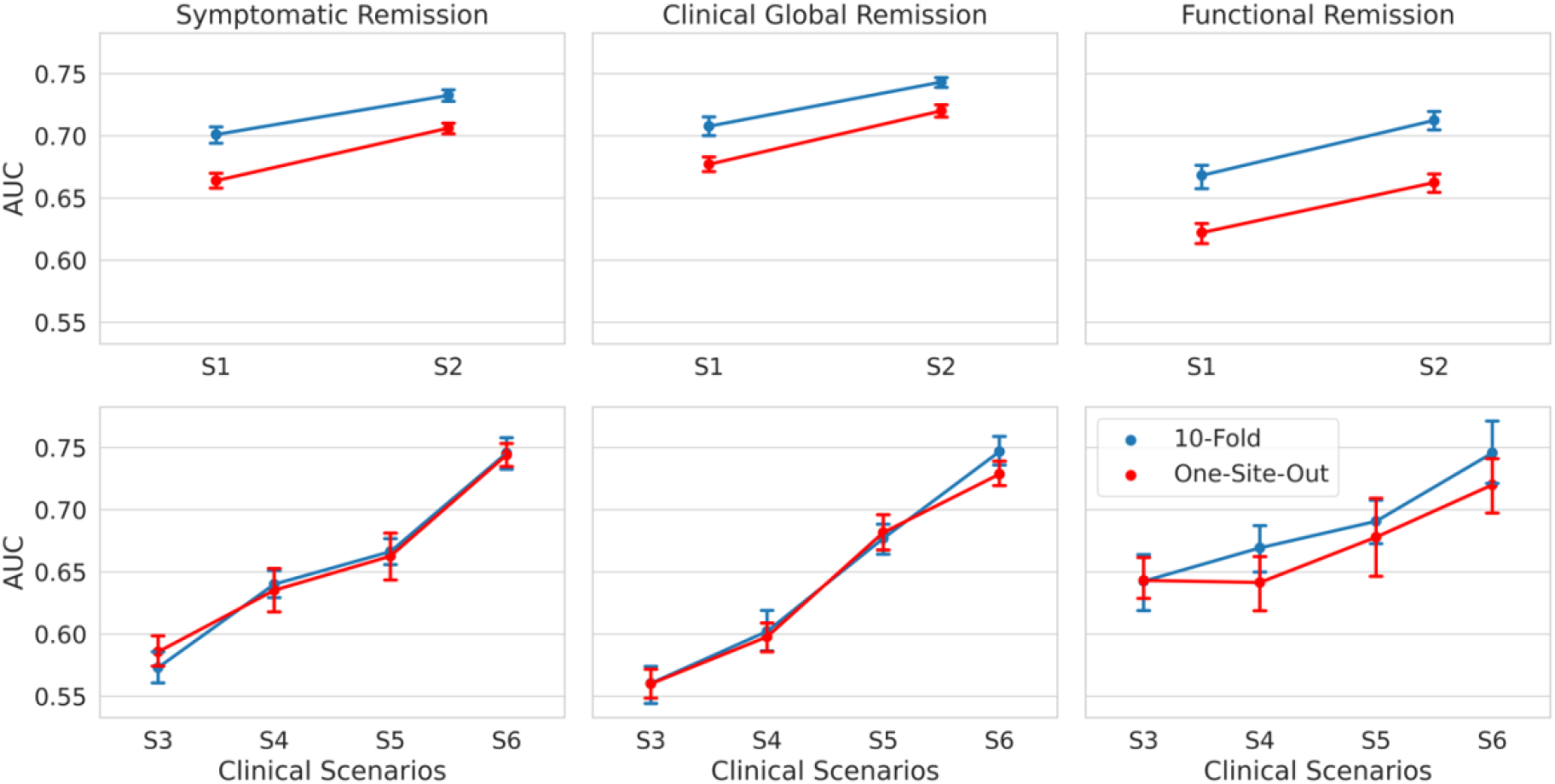
AUCs of the model across three outcome measures (first column: symptomatic remission, second column: clinical global remission, and third column: functional remission) for six clinical scenarios. The x-axes represent the clinical scenarios in Phase 1 (S_1_ and S_2_) and Phase 2 of the study (S_3_, S_4_, S_5_, and S_6_). The y-axis shows the AUC. The blue and red lines represent the results for 10-fold and one-site-out cross-validation, respectively. The error bars show the standard deviation of performance across 20 repetitions. The results in the first row show the AUCs in phase 1 in a 4-week prediction. The added use of time point W_1_ increases the AUC for all outcome measures, with added AUC ranging from 0.03 to 0.05. The second row shows the results of phase 2 in a 10-week prediction. Except for one instance (input: W_0_-W_1_, outcome: functional remission; validation: one-site out), each added time point further increases the prediction performance for all outcome measures.

**Table 2.**
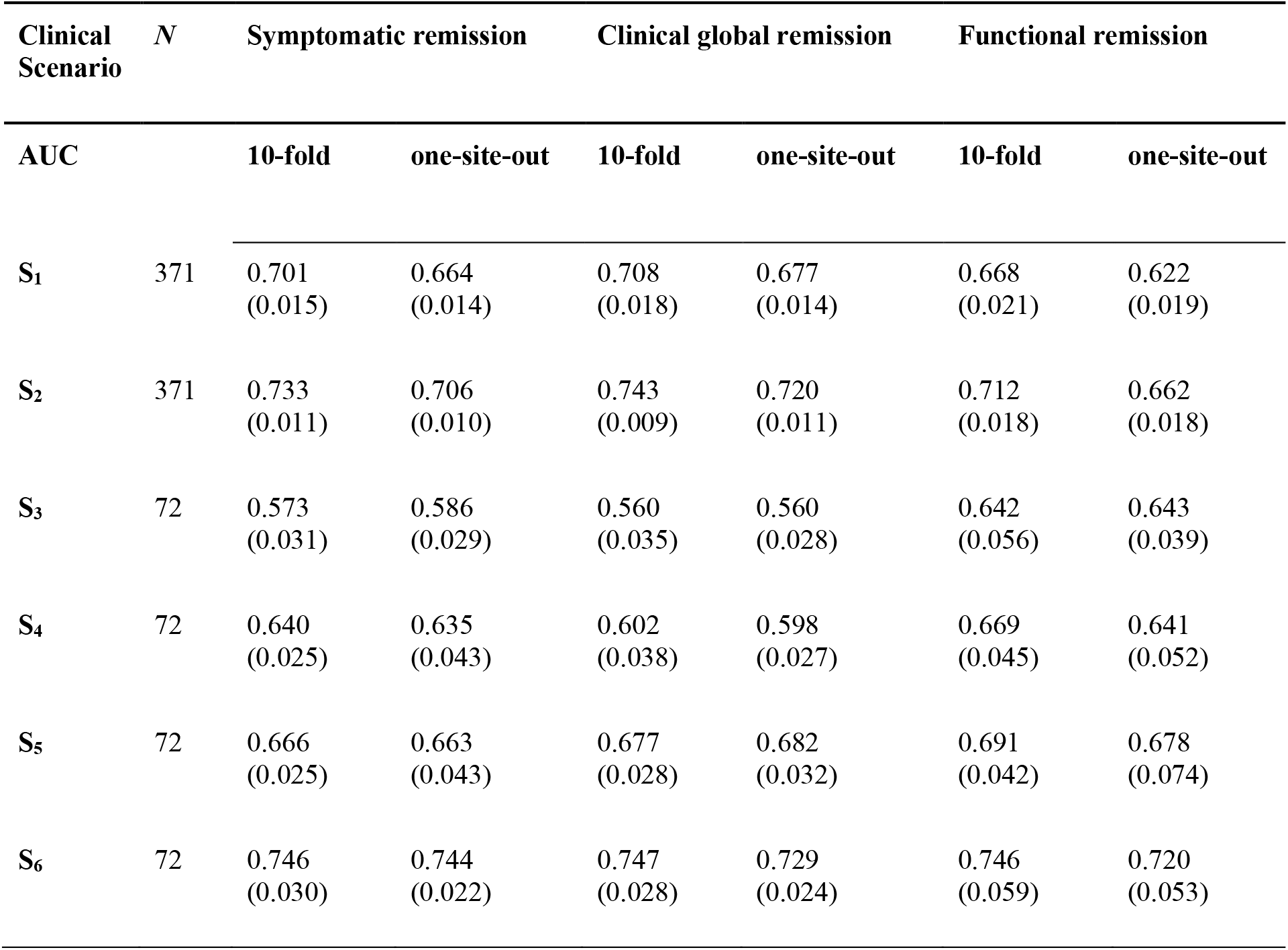
Performance of the prediction models predicting three outcome measures (symptomatic remission, clinical global remission and functional remission) for six clinical scenarios (S_1_-S_6_). Performance is measured by the area under the receiver operating characteristic curve (AUC). The values are averaged over 20 repetitions of 10-fold and one-site-out cross-validation. The values in the parentheses represent the standard deviation over these repetitions.

| Clinical Scenario | $N$ | Symptomatic remission | | Clinical global remission | | Functional remission | |
| --- | --- | --- | --- | --- | --- | --- | --- |
|  |  | 10-fold | one-site-out | 10-fold | one-site-out | 10-fold | one-site-out |
| $S_1$ | 371 | 0.701<br>(0.015) | 0.664<br>(0.014) | 0.708<br>(0.018) | 0.677<br>(0.014) | 0.668<br>(0.021) | 0.622<br>(0.019) |
| $S_2$ | 371 | 0.733<br>(0.011) | 0.706<br>(0.010) | 0.743<br>(0.009) | 0.720<br>(0.011) | 0.712<br>(0.018) | 0.662<br>(0.018) |
| $S_3$ | 72 | 0.573<br>(0.031) | 0.586<br>(0.029) | 0.560<br>(0.035) | 0.560<br>(0.028) | 0.642<br>(0.056) | 0.643<br>(0.039) |
| $S_4$ | 72 | 0.640<br>(0.025) | 0.635<br>(0.043) | 0.602<br>(0.038) | 0.598<br>(0.027) | 0.669<br>(0.045) | 0.641<br>(0.052) |
| $S_5$ | 72 | 0.666<br>(0.025) | 0.663<br>(0.043) | 0.677<br>(0.028) | 0.682<br>(0.032) | 0.691<br>(0.042) | 0.678<br>(0.074) |
| $S_6$ | 72 | 0.746<br>(0.030) | 0.744<br>(0.022) | 0.747<br>(0.028) | 0.729<br>(0.024) | 0.746<br>(0.059) | 0.720<br>(0.053) |

### Incorporating model uncertainty reduces the risk of decision making

To quantitatively evaluate the advantage of using uncertainty-aware predictions, we evaluated the accuracy of the symptomatic remission prediction model in six clinical scenarios at four levels of conservativeness:

● Level 0, in which the trivial threshold-based approach is used for decision-making. A hard threshold of 0.5 is applied to the predicted probability of remission to decide between non-remission (below the threshold) or remission (equal or above the threshold) decisions. In fact, the proposed decision-making method is not used.
● Level 1, in which the clinician abstains from utilizing the model’s predictions that lie in the ‘unsure (US)’ category (when the model says “I do not know”).
● Level 2 of conservativeness, where predictions from the three most uncertain prediction categories (US, UR, and UN) are not used for clinical decision-making.
● Level 3, wherein the most conservative usage of model predictions only the most certain decisions of the model (the DR and DN categories) are employed by the clinicians for decision-making.

The results of applying increasing levels of conservativeness are presented in Table 3. An incremental trend in the accuracy of decisions is seen, when rising the conservativeness level from 0 to 3 (see also Fig. 8), which is, naturally, accompanied by a decrease in the number of patients for whom an ML-aided decision is made (represented by decisiveness in the table). At level 1, and by excluding ∼10% of decisions in the US category, the accuracy of the model is improved by ∼0.06 across all clinical scenarios. At level 2, excluding ∼50% (the uncertain predictions in US, UR, and UN) from decision-making, results in a further increase in accuracy to ∼0.86. By restricting the decision-making to DR and DN categories at level 3, the accuracy of the model is increased to ∼0.95 within ∼16% of patients with decisions. Thus, the clinicians can trust the DR and DN decisions with 0.95 confidence (although without being able to use decisions for ∼84% of their patients). This is a crucial feature for more trustworthy decision-making in clinics because the users (i.e., clinicians) not only receive an ML-aided data-driven recommendation from the machine but are also informed about the risk involved in relying on these predictions. The more confidence in the model’s predictions (in DR and DN categories), the less risk is involved in AI-aided decision-making.

**Figure 8.**
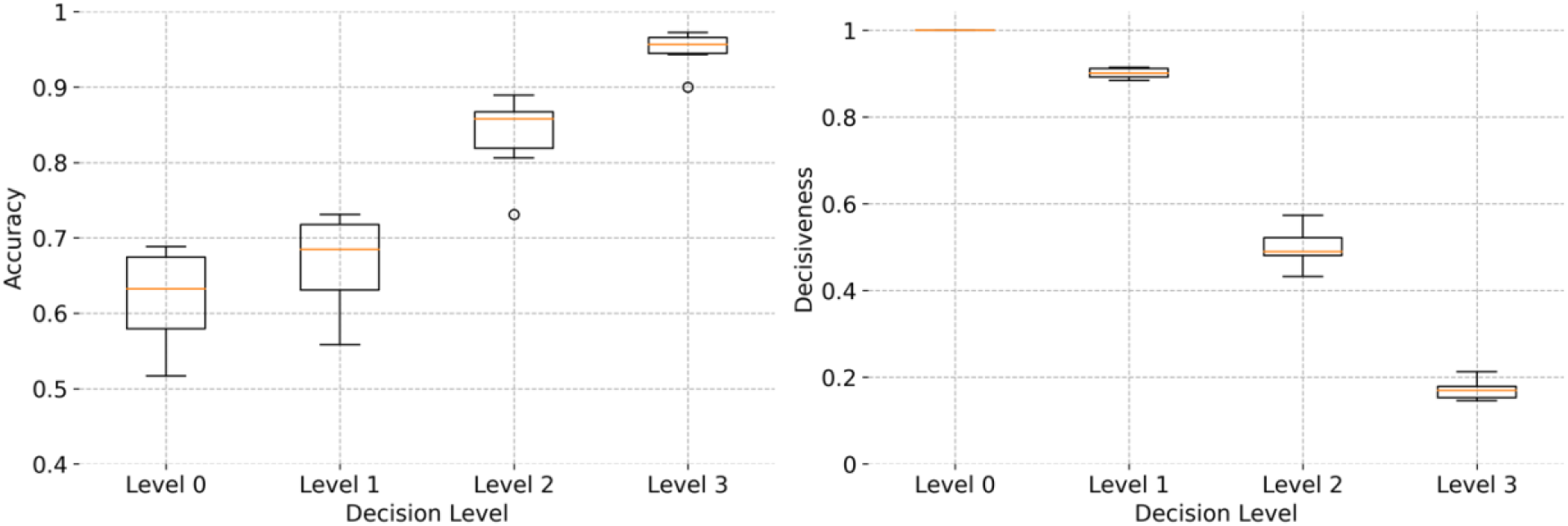
A comparison between the accuracy and decisiveness of ML-aided decision-making at four levels of conservativeness (0-3). The boxplots represent the average accuracy (left) and decisiveness (right) over 20 repetitions of 10-fold cross-validation across six clinical scenarios. Using more certain predictions for decision-making results in less decisive but more accurate models.

**Table 3.**
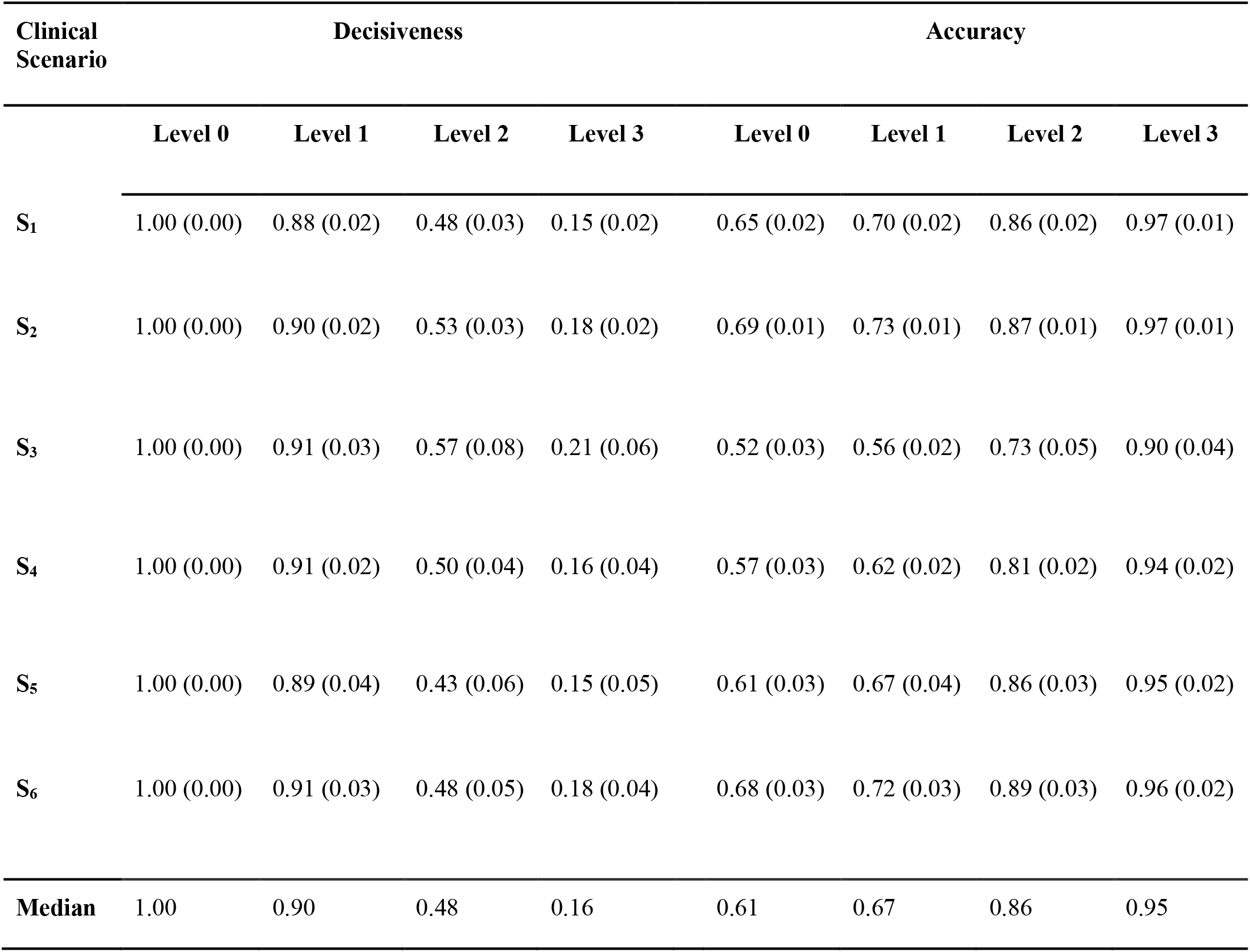
The decisiveness (the proportion of decided sample to total sample) and the accuracy of decision for symptomatic remission, for six clinical scenarios (rows) and four decision levels (columns). The values are averaged over 20 repetitions of 10-fold cross-validation. The values in the parentheses represent the standard deviation over these repetitions.

| Clinical Scenario | Decisiveness |  |  |  | Accuracy |  |  |  |
| --- | --- | --- | --- | --- | --- | --- | --- | --- |
|  | Level 0 | Level 1 | Level 2 | Level 3 | Level 0 | Level 1 | Level 2 | Level 3 |
| **S<sub>1</sub>** | 1.00 (0.00) | 0.88 (0.02) | 0.48 (0.03) | 0.15 (0.02) | 0.65 (0.02) | 0.70 (0.02) | 0.86 (0.02) | 0.97 (0.01) |
| **S<sub>2</sub>** | 1.00 (0.00) | 0.90 (0.02) | 0.53 (0.03) | 0.18 (0.02) | 0.69 (0.01) | 0.73 (0.01) | 0.87 (0.01) | 0.97 (0.01) |
| **S<sub>3</sub>** | 1.00 (0.00) | 0.91 (0.03) | 0.57 (0.08) | 0.21 (0.06) | 0.52 (0.03) | 0.56 (0.02) | 0.73 (0.05) | 0.90 (0.04) |
| **S<sub>4</sub>** | 1.00 (0.00) | 0.91 (0.02) | 0.50 (0.04) | 0.16 (0.04) | 0.57 (0.03) | 0.62 (0.02) | 0.81 (0.02) | 0.94 (0.02) |
| **S<sub>5</sub>** | 1.00 (0.00) | 0.89 (0.04) | 0.43 (0.06) | 0.15 (0.05) | 0.61 (0.03) | 0.67 (0.04) | 0.86 (0.03) | 0.95 (0.02) |
| **S<sub>6</sub>** | 1.00 (0.00) | 0.91 (0.03) | 0.48 (0.05) | 0.18 (0.04) | 0.68 (0.03) | 0.72 (0.03) | 0.89 (0.03) | 0.96 (0.02) |
| **Median** | 1.00 | 0.90 | 0.48 | 0.16 | 0.61 | 0.67 | 0.86 | 0.95 |

## Discussion

This study set out to build a prediction model that has the potential to be used as a tool to assist in clinical decision-making. To the best of our knowledge, we are the first to build a model that fulfills three crucial criteria for use in clinical practice. Firstly, by using the LSTM architecture, the model was trained on time series data. Previous studies^4–8^, using different architectures, only used baseline data in their prediction models. These architectures do not accommodate the use of time series data in the same prediction model. The LSTM architecture is flexible; it is possible to build one model that can be used at different points in time, adding the additional data that is available at a specific moment. Therefore, unlike other prediction architectures, it fits real-world data^35^, where a patient is assessed regularly.

Secondly, our architecture is multi-task, so one prediction model can predict multiple outcome measures simultaneously. This feature has been highlighted as important by our panel of advisors which regularly contributes to the understanding of real-world patient and doctor needs. The involvement of such panels was found to be crucial in building trust in AI solutions in healthcare, among both patients and doctors^36^. In previous studies, when predicting multiple outcome measures, separate prediction models were needed, one for each outcome measure;^4–7, 37–44^. In this study, we were able to predict symptomatic, clinical global and functional remission in just one model. The multi-task model provides a way for clinicians to aim for recovery in several domains.

Thirdly, we used the uncertainty of predictions to adjust the prediction accuracy and build a novel decision-making module for more individualized decision-making. Thus, clinicians can say they are sure or not sure about an individual prediction if the model is certain or uncertain respectively, which reduces the chance of making a wrong treatment decision. Also, this is an important feature for machine learning models to be considered trustworthy by clinicians and patients alike^17^. Using this flexible multi-task architecture that incorporates the uncertainty of individual predictions, we took a leap forward toward improving patient care with the help of machine learning prediction models.

Considering the specific characteristics of our current solution, we found accuracies of up to 0.72 AUC for 4-week prediction and up to 0.74 AUC for 10-week prediction using one-site-out as a validation method. These results are comparable to previously conducted studies. We have shown that the use of multiple time points increased the accuracy of prediction for all outcome measures for both 4-week and 10-week predictions.

Although the accuracy of our models is not above the 80% threshold suggested by the APA^45^ when all patients are incorporated, we still consider our models to be clinically relevant, as they provide a certainty measure for each prediction. Besides, when the uncertain predictions are discarded (our ‘decision-making level 2’), across the six different prediction models 43–57% of the patient sample was still included, with accuracies ranging from 0.73 to 0.89, with five of our six models achieving an accuracy above 0.8. This is why we consider this feature an important step toward reaching our goal of building an interactive tool for individual psychosis prognosis prediction.

In other fields of healthcare, comparable approaches oriented towards computing disease risk have been developed^46, 47^, albeit without the use of a certainty measure for each prediction. However, we consider this an indispensable feature, given the potentially severe consequences of wrong decision-making in treatment. For psychosis treatment specifically, unnecessary side-effects of antipsychotic medication or longer duration of untreated psychosis are to be considered in this aspect.

## Limitations

Our LSTM model can use time series data, but currently, this is only possible when data from all previous time points are also available. In clinical practice, this could be a potential problem, in situations where a patient misses an appointment or is not capable of providing information in certain exams or questionnaires at some point. This problem can be solved by using LSTM models that can handle missing measurements^48^ or by incorporating the length of time intervals in the modeling process^49^. We consider these as possible future directions to extend our work.

Considering the data the model was tested on, all patients in the dataset used amisulpride in the first phase, and amisulpride or olanzapine in the second phase of their treatment. Therefore our model only applies to patients using amisulpride. Also, not all potentially relevant predictor variables were available in our data set, such as childhood adverse events. A larger dataset with a more diverse and heterogeneous sample would improve the clinical applicability of future models.

## Conclusion

In conclusion, we developed and tested a psychosis prognosis prediction model that has properties that are required for use in clinical practice. Using a flexible multi-task LSTM architecture that was optimized for this goal, the ability to use time series data was shown to be of great importance once prediction models will be used in clinical care. By building a multi-task model, different clinically relevant outcomes can be predicted simultaneously. For more reliable decision-making, we built a decision-making module that considers the uncertainty of individual predictions and we demonstrated its usefulness.

## Data Availability

All data produced in the present study are available upon reasonable request to the authors.

## Acknowledgments

This work was supported by ZonMw (project ID 63631 0011) and by a research grant from the AI for Health working group of the TU/e-WUR-UU-UMCU (EWUU) alliance.

## SUPPLEMENTARY MATERIALS

**sTable 1.**
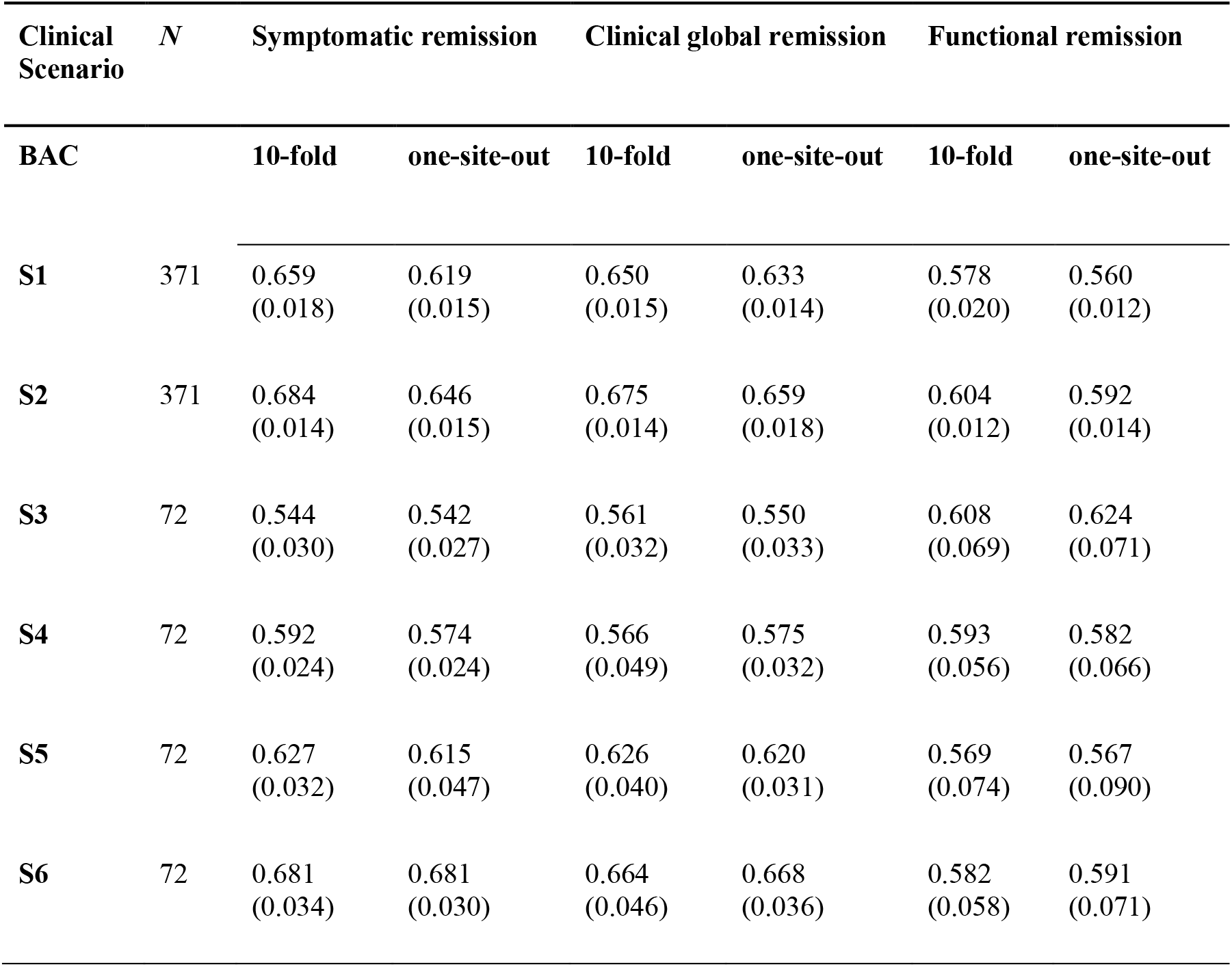
Balanced accuracy (BAC) of the prediction models predicting three outcome measures (symptomatic remission, clinical global remission and functional remission) for six clinical scenarios (S_1_-S_6_). The values are averaged over 20 repetitions of 10-fold and one-site-out cross-validation. The values in the parentheses represent the standard deviation over these repetitions.

**sTable 2.** Sensitivity of the prediction models predicting three outcome measures (symptomatic remission, clinical global remission and functional remission) for six clinical scenarios (S_1_-S_6_). The values are averaged over 20 repetitions of 10-fold and one-site-out cross-validation. The values in the parentheses represent the standard deviation over these repetitions.

| SENSITIVITY |  | 10-fold | one-site-out | 10-fold | one-site-out | 10-fold | one-site-out |
| --- | --- | --- | --- | --- | --- | --- | --- |
| **S1** | 371 | 0.628 | 0.584 | 0.495 | 0.498 | 0.283 | 0.272 |
|  |  | (0.032) | (0.033) | (0.032) | (0.032) | (0.041) | (0.031) |
| **S2** | 371 | 0.688 | 0.660 | 0.574 | 0.578 | 0.352 | 0.342 |
|  |  | (0.023) | (0.023) | (0.033) | (0.038) | (0.034) | (0.030) |
| **S3** | 72 | 0.769 | 0.836 | 0.585 | 0.631 | 0.438 | 0.506 |
|  |  | (0.068) | (0.065) | (0.064) | (0.068) | (0.138) | (0.149) |
| **S4** | 72 | 0.772 | 0.824 | 0.529 | 0.579 | 0.375 | 0.388 |
|  |  | (0.074) | (0.049) | (0.092) | (0.071) | (0.107) | (0.134) |
| **S5** | 72 | 0.703 | 0.748 | 0.531 | 0.567 | 0.256 | 0.281 |
|  |  | (0.071) | (0.090) | (0.080) | (0.068) | (0.143) | (0.185) |
| **S6** | 72 | 0.683 | 0.757 | 0.556 | 0.598 | 0.250 | 0.281 |
|  |  | (0.072) | (0.064) | (0.089) | (0.073) | (0.128) | (0.156) |

**sTable 3.** Sensitivity of the prediction models predicting three outcome measures (symptomatic remission, clinical global remission and functional remission) for six clinical scenarios (S_1_-S_6_). The values are averaged over 20 repetitions of 10-fold and one-site-out cross-validation. The values in the parentheses represent the standard deviation over these repetitions.

| SPECIFICITY |  | 10-fold | one-site-out | 10-fold | one-site-out | 10-fold | one-site-out |
| --- | --- | --- | --- | --- | --- | --- | --- |
| **S1** | 371 | 0.690 | 0.653 | 0.804 | 0.768 | 0.873 | 0.848 |
|  |  | (0.044) | (0.034) | (0.022) | (0.026) | (0.015) | (0.023) |
| **S2** | 371 | 0.680 | 0.633 | 0.777 | 0.740 | 0.857 | 0.842 |
|  |  | (0.029) | (0.026) | (0.034) | (0.026) | (0.018) | (0.015) |
| **S3** | 72 | 0.319 | 0.247 | 0.536 | 0.469 | 0.779 | 0.742 |
|  |  | (0.062) | (0.092) | (0.063) | (0.058) | (0.035) | (0.044) |
| **S4** | 72 | 0.412 | 0.323 | 0.604 | 0.571 | 0.811 | 0.776 |
|  |  | (0.084) | (0.047) | (0.055) | (0.051) | (0.041) | (0.037) |
| **S5** | 72 | 0.550 | 0.482 | 0.720 | 0.673 | 0.882 | 0.853 |
|  |  | (0.072) | (0.085) | (0.060) | (0.048) | (0.032) | (0.033) |
| **S6** | 72 | 0.678 | 0.605 | 0.771 | 0.738 | 0.914 | 0.900 |
|  |  | (0.035) | (0.061) | (0.035) | (0.036) | (0.029) | (0.032) |

**sFigure 1.**
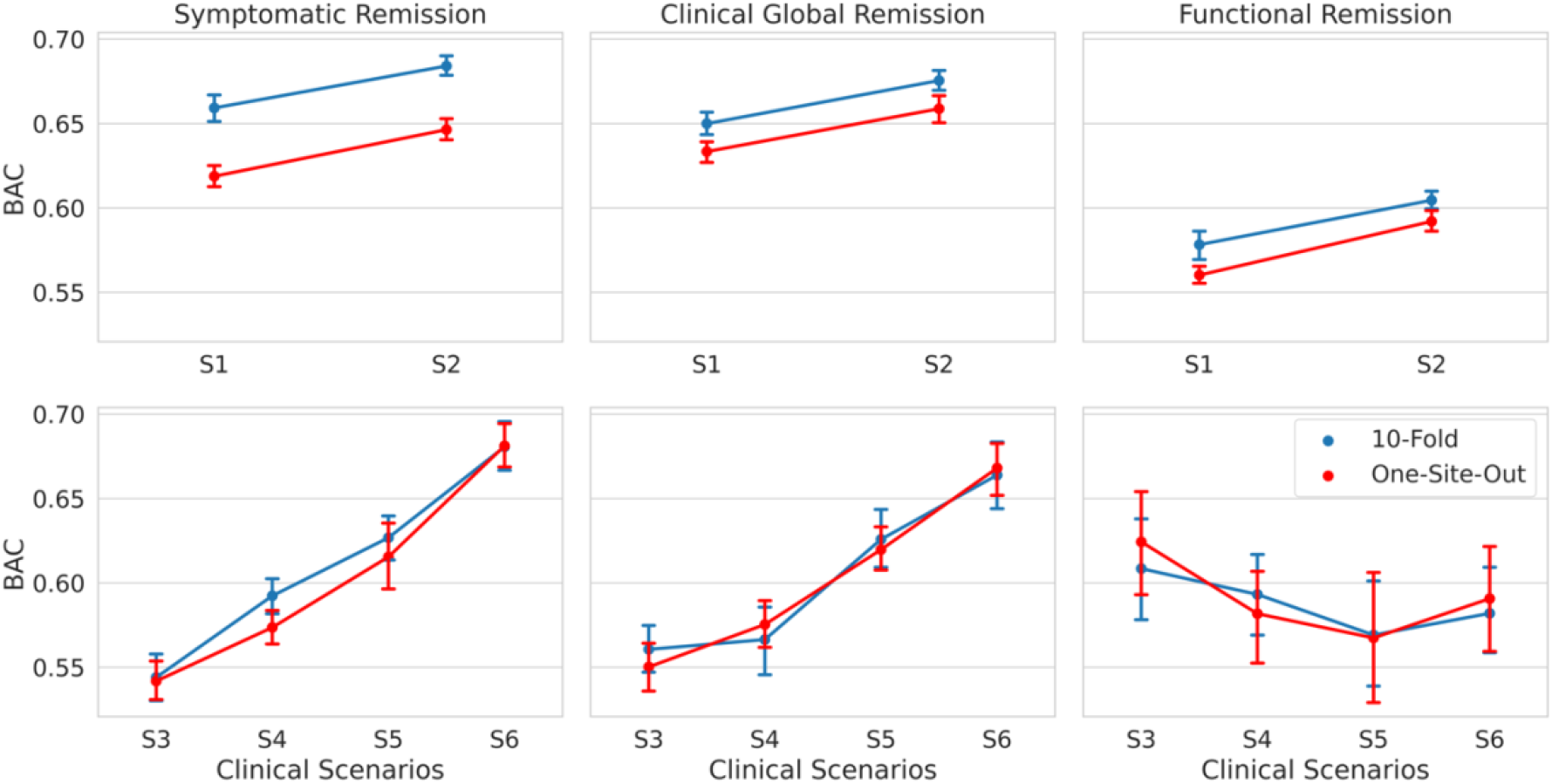
Balanced accuracies (BACs) of the model across three outcome measures (first column: symptomatic remission, second column: clinical global remission, and third column: functional remission) for six clinical scenarios. The x-axes represent the clinical scenarios in Phase 1 (S1 and S2) and Phase 2 of the study (S3, S4, S5, and S6). The y-axis shows the BAC. The blue and red lines represent the results for 10-fold and one-site-out cross-validation, respectively. The error bars show the standard deviation of performance across 20 repetitions. The results in the first row show the BAC in phase 1 in a 4-week prediction. The added use of time point W_1_ increases the BAC for all outcome measures. This is mainly due to the increased sensitivity of the model when a new time point is added. The second row shows the results of phase 2 in a 10-week prediction. Except for one instance (functional remission), each added time point further increases the BAC for all outcome measures. The Increase in the BACs in this case is a byproduct of the increased specificity (see sFigure 2 and sFigure 3) of the model when a new time point is added.

**sFigure 2.**
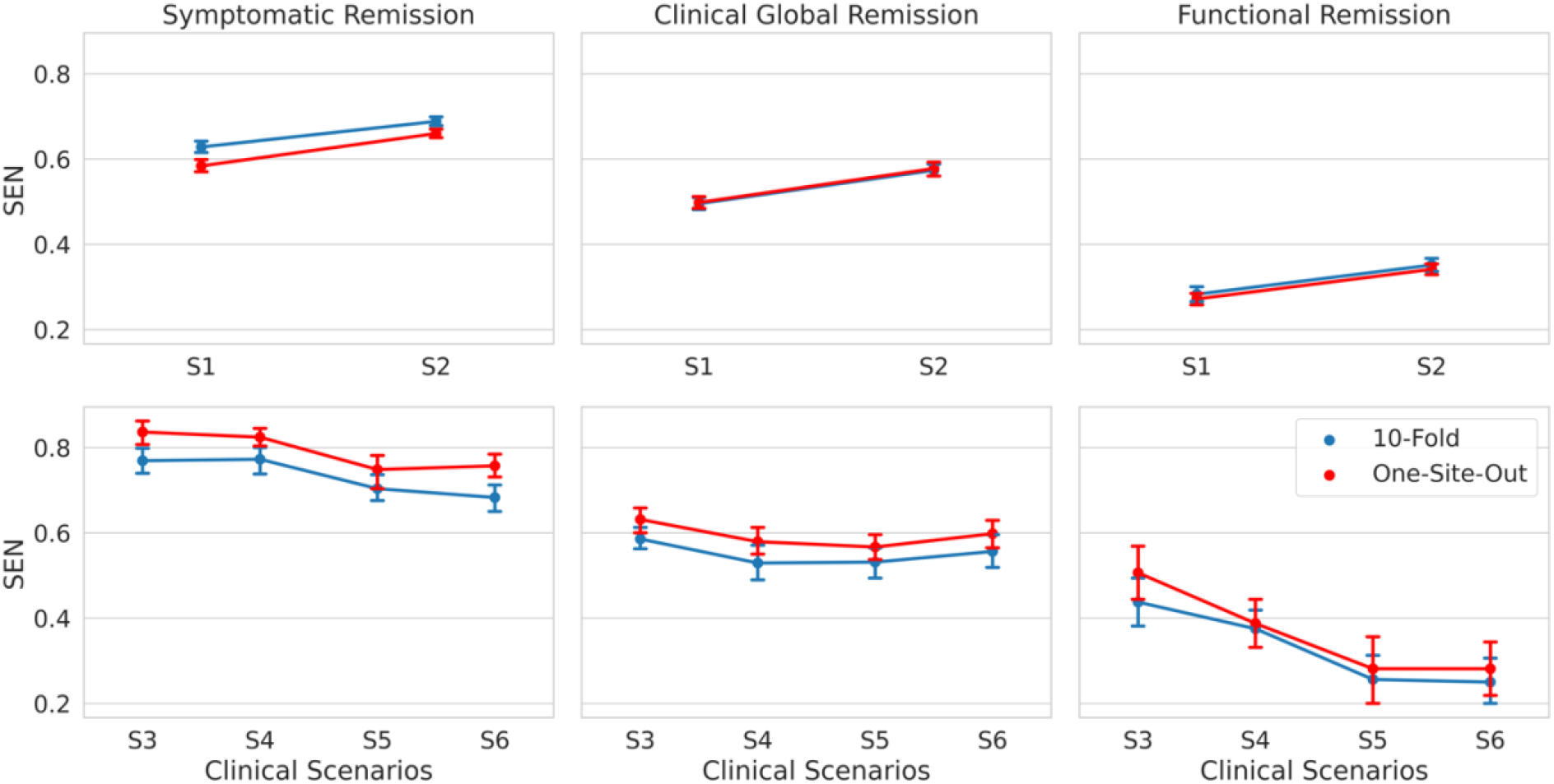
Sensitivity (SEN) of the model across three outcome measures (first column: symptomatic remission, second column: clinical global remission, and third column: functional remission) for six clinical scenarios. The x-axes represent the clinical scenarios in Phase 1 (S1 and S2) and Phase 2 of the study (S3, S4, S5, and S6). The y-axis shows the SEN. The blue and red lines represent the results for 10-fold and one-site-out cross-validation, respectively. The error bars show the standard deviation of SENs across 20 repetitions. The results in the first row show the SEN in phase 1 in a 4-week prediction. The added use of time point W_1_ increases the SEN for all outcome measures. The second row shows the results of phase 2 in a 10-week prediction. In most cases adding a new time point results in a reduced sensitivity of the model.

**sFigure 3.**
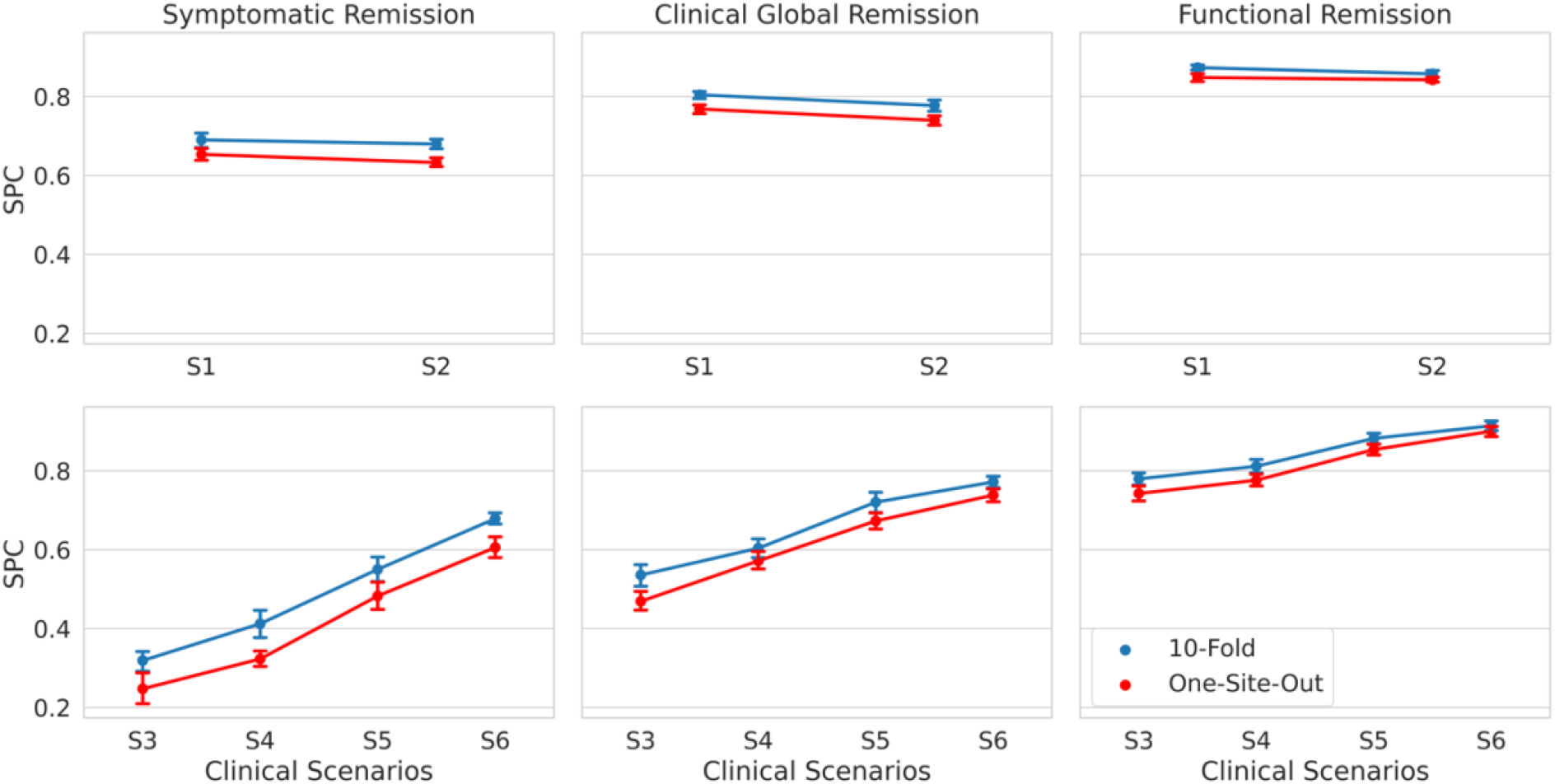
Specificity (SPC) of the model across three outcome measures (first column: symptomatic remission, second column: clinical global remission, and third column: functional remission) for six clinical scenarios. The x-axes represent the clinical scenarios in Phase 1 (S1 and S2) and Phase 2 of the study (S3, S4, S5, and S6). The y-axis shows the SPC. The blue and red lines represent the results for 10-fold and one-site-out cross-validation, respectively. The error bars show the standard deviation of SPCs across 20 repetitions. The results in the first row show the SPC in phase 1 in a 4-week prediction. The added use of time point W_1_ slightly decreases the SPC for all outcome measures. The second row shows the results of phase 2 in a 10-week prediction. In all cases adding a new time point results in higher model specificity.

## References

1. Bzdok D, Meyer-Lindenberg A. Machine Learning for Precision Psychiatry: Opportunities and Challenges. Biol Psychiatry Cogn Neurosci Neuroimaging. 2018;3(3):223–230. doi:10.1016/j.bpsc.2017.11.007

2. Chekroud AM, Bondar J, Delgadillo J, et al. The promise of machine learning in predicting treatment outcomes in psychiatry. World Psychiatry. 2021;20(2):154–170. doi:10.1002/wps.20882

3. Salazar de Pablo G, Studerus E, Vaquerizo-Serrano J, et al. Implementing Precision Psychiatry: A Systematic Review of Individualized Prediction Models for Clinical Practice. Schizophr Bull. 2021;47(2):284–297. doi:10.1093/schbul/sbaa120

4. Koutsouleris N, Kahn RS, Chekroud AM, et al. Multisite prediction of 4-week and 52-week treatment outcomes in patients with first-episode psychosis: a machine learning approach. Lancet Psychiatry. 2016;3(10):935–946. doi:10.1016/S2215-0366(16)30171-7

5. Fond G, Bulzacka E, Boucekine M, et al. Machine learning for predicting psychotic relapse at 2 years in schizophrenia in the national FACE-SZ cohort. Prog Neuropsychopharmacol Biol Psychiatry. 2019;92:8–18. doi:10.1016/j.pnpbp.2018.12.005

6. Leighton SP, Krishnadas R, Chung K, et al. Predicting one-year outcome in first episode psychosis using machine learning. PLoS One. 2019;14(3):e0212846. doi:10.1371/journal.pone.0212846

7. de Nijs J, Burger TJ, Janssen RJ, et al. Individualized prediction of three-and six-year outcomes of psychosis in a longitudinal multicenter study: a machine learning approach. NPJ Schizophr. 2021;7(1):34. doi:10.1038/s41537-021-00162-3

8. Austin JC, Hippman C, Honer WG. Descriptive and numeric estimation of risk for psychotic disorders among affected individuals and relatives: implications for clinical practice. Psychiatry Res. 2012;196(1):52–56. doi:10.1016/j.psychres.2012.02.005

9. Soldatos RF, Cearns M, Nielsen MØ, et al. Prediction of Early Symptom Remission in Two Independent Samples of First-Episode Psychosis Patients Using Machine Learning. Schizophr Bull. 2022;48(1):122–133. doi:10.1093/schbul/sbab107

10. Lin E, Lin CH, Lane HY. Applying a bagging ensemble machine learning approach to predict functional outcome of schizophrenia with clinical symptoms and cognitive functions. Sci Rep. 2021;11(1):6922. doi:10.1038/s41598-021-86382-0

11. Li Y, Zhang L, Zhang Y, et al. A Random Forest Model for Predicting Social Functional Improvement in Chinese Patients with Schizophrenia After 3 Months of Atypical Antipsychotic Monopharmacy: A Cohort Study. Neuropsychiatr Dis Treat. 2021;17:847–857. doi:10.2147/NDT.S280757

12. Leighton SP, Upthegrove R, Krishnadas R, et al. Development and validation of multivariable prediction models of remission, recovery, and quality of life outcomes in people with first episode psychosis: a machine learning approach. Lancet Digit Health. 2019;1(6):e261–e270. doi:10.1016/S2589-7500(19)30121-9

13. Basaraba CN, Scodes JM, Dambreville R, et al. Prediction Tool for Individual Outcome Trajectories Across the Next Year in First-Episode Psychosis in Coordinated Specialty Care. JAMA Psychiatry. 2023;80(1):49–56. doi:10.1001/jamapsychiatry.2022.3571

14. Caruana R. Multitask Learning. In: Thrun S, Pratt L., eds. Learning to Learn. Springer US; 1998:95–133. doi:10.1007/978-1-4615-5529-2_5

15. Kaushik S, Choudhury A, Sheron PK, et al. AI in Healthcare: Time-Series Forecasting Using Statistical, Neural, and Ensemble Architectures. Front Big Data. 2020;3:4. doi:10.3389/fdata.2020.00004

16. Kendall A, Gal Y. What uncertainties do we need in Bayesian deep learning for computer vision? arXiv *[csCV]*. Published online March 15, 2017. Accessed December 9, 2022. https://proceedings.neurips.cc/paper/2017/hash/2650d6089a6d640c5e85b2b88265dc2b-Abstract.html

17. Kompa B, Snoek J, Beam AL. Second opinion needed: communicating uncertainty in medical machine learning. NPJ Digit Med. 2021;4(1):4. doi:10.1038/s41746-020-00367-3

18. Meijerink L, Cinà G, Tonutti M. Uncertainty estimation for classification and risk prediction on medical tabular data. arXiv *[statML]*. Published online April 13, 2020. http://arxiv.org/abs/2004.05824

19. Kahn RS, Winter van Rossum I, Leucht S, et al. Amisulpride and olanzapine followed by open-label treatment with clozapine in first-episode schizophrenia and schizophreniform disorder (OPTiMiSE): a three-phase switching study. Lancet Psychiatry. 2018;5(10):797–807. doi:10.1016/S2215-0366(18)30252-9

20. Andreasen NC, Carpenter WT Jr, Kane JM, Lasser RA, Marder SR, Weinberger DR. Remission in schizophrenia: proposed criteria and rationale for consensus. Am J Psychiatry. 2005;162(3):441–449. doi:10.1176/appi.ajp.162.3.441

21. Kay SR, Fiszbein A, Opler LA. The positive and negative syndrome scale (PANSS) for schizophrenia. Schizophr Bull. 1987;13(2):261–276. doi:10.1093/schbul/13.2.261

22. Guy W. *ECDEU Assessment Manual for Psychopharmacology:* 1976. National Institute of Mental Health; 1976.

23. Morosini PL, Magliano L, Brambilla L, Ugolini S, Pioli R. Development, reliability and acceptability of a new version of the DSM-IV Social and Occupational Functioning Assessment Scale (SOFAS) to assess routine social functioning. Acta Psychiatr Scand. 2000;101(4):323–329. https://www.ncbi.nlm.nih.gov/pubmed/10782554

24. Hochreiter S, Schmidhuber J. Long short-term memory. Neural Comput. 1997;9(8):1735– 1780. doi:10.1162/neco.1997.9.8.1735

25. Lindemann B, Müller T, Vietz H, Jazdi N, Weyrich M. A survey on long short-term memory networks for time series prediction. Procedia CIRP. 2021;99:650–655. doi:10.1016/j.procir.2021.03.088

26. Fischer T, Krauss C. Deep learning with long short-term memory networks for financial market predictions. Eur J Oper Res. 2018;270(2):654–669. doi:10.1016/j.ejor.2017.11.054

27. Cox DR. Principles of Statistical Inference. Cambridge University Press; 2006. https://play.google.com/store/books/details?id=nRgtGZXi2KkC

28. Zadeh LA. Fuzzy logic. Computer. 1988;21(4):83–93. doi:10.1109/2.53

29. Mamdani EH. Application of fuzzy algorithms for control of simple dynamic plant. Proceedings of the Institution of Electrical Engineers. 1974;121(12):1585–1588. doi:10.1049/piee.1974.0328

30. Srivastava N, Hinton G, Krizhevsky A, Sutskever I, Salakhutdinov R. Dropout: A simple way to prevent neural networks from overfitting. Accessed December 16, 2022. https://www.jmlr.org/papers/volume15/srivastava14a/srivastava14a.pdf?utm_content=buffer79b43&utm_medium=social&utm_source=twitter.com&utm_campaign=buffer,

31. Gal Y, Ghahramani Z. Dropout as a Bayesian Approximation: Representing Model Uncertainty in Deep Learning. In: Balcan MF, Weinberger KQ, eds. Proceedings of The 33rd International Conference on Machine Learning. Vol 48. Proceedings of Machine Learning Research. PMLR; 20--22 Jun 2016:1050–1059. https://proceedings.mlr.press/v48/gal16.html

32. Nixon J, Dusenberry M, Jerfel G, et al. Measuring calibration in deep learning. arXiv [csLG]. Published online April 2, 2019. Accessed December 9, 2022. http://openaccess.thecvf.com/content_CVPRW_2019/papers/Uncertainty%20and%20Robustness%20in%20Deep%20Visual%20Learning/Nixon_Measuring_Calibration_in_Deep_Learning_CVPRW_2019_paper.pdf

33. Van Calster B, McLernon DJ, van Smeden M, Wynants L, Steyerberg EW, Topic Group “Evaluating diagnostic tests and prediction models” of the STRATOS initiative. Calibration: the Achilles heel of predictive analytics. BMC Med. 2019;17(1):230. doi:10.1186/s12916-019-1466-7

34. van Smeden M, Reitsma JB, Riley RD, Collins GS, Moons KG. Clinical prediction models: diagnosis versus prognosis. J Clin Epidemiol. 2021;132:142–145. doi:10.1016/j.jclinepi.2021.01.009

35. Makady A, de Boer A, Hillege H, Klungel O, Goettsch W, (on behalf of GetReal Work Package 1). What Is Real-World Data? A Review of Definitions Based on Literature and Stakeholder Interviews. Value Health. 2017;20(7):858–865. doi:10.1016/j.jval.2017.03.008

36. Banerjee S, Alsop P, Jones L, Cardinal RN. Patient and public involvement to build trust in artificial intelligence: A framework, tools, and case studies. Patterns (N Y). 2022;3(6):100506. doi:10.1016/j.patter.2022.100506

37. Albert N, Bertelsen M, Thorup A, et al. Predictors of recovery from psychosis Analyses of clinical and social factors associated with recovery among patients with first-episode psychosis after 5 years. Schizophr Res. 2011;125(2-3):257–266. doi:10.1016/j.schres.2010.10.013

38. de Wit S, Ziermans TB, Nieuwenhuis M, et al. Individual prediction of long-term outcome in adolescents at ultra-high risk for psychosis: Applying machine learning techniques to brain imaging data. Hum Brain Mapp. 2017;38(2):704–714. doi:10.1002/hbm.23410

39. Gasquet I, Haro JM, Tcherny-Lessenot S, Chartier F, Lépine JP. Remission in the outpatient care of schizophrenia: 3-year results from the Schizophrenia Outpatients Health Outcomes (SOHO) study in France. Eur Psychiatry. 2008;23(7):491–496. doi:10.1016/j.eurpsy.2008.03.012

40. Koutsouleris N, Kambeitz-Ilankovic L, Ruhrmann S, et al. Prediction Models of Functional Outcomes for Individuals in the Clinical High-Risk State for Psychosis or With Recent-Onset Depression: A Multimodal, Multisite Machine Learning Analysis. JAMA Psychiatry. 2018;75(11):1156–1172. doi:10.1001/jamapsychiatry.2018.2165

41. Lambert M, Karow A, Leucht S, Schimmelmann BG, Naber D. Remission in schizophrenia: validity, frequency, predictors, and patients’ perspective 5 years later. Dialogues Clin Neurosci. 2010;12(3):393–407. doi:10.31887/DCNS.2010.12.3/mlambert

42. Lambert M, Schimmelmann BG, Naber D, et al. Prediction of remission as a combination of symptomatic and functional remission and adequate subjective well-being in 2960 patients with schizophrenia. J Clin Psychiatry. 2006;67(11):1690–1697. doi:10.4088/jcp.v67n1104

43. Malla A, Norman R, Schmitz N, et al. Predictors of rate and time to remission in first-episode psychosis: a two-year outcome study. Psychol Med. 2006;36(5):649–658. doi:10.1017/S0033291706007379

44. Caton CLM, Hasin DS, Shrout PE, et al. Predictors of psychosis remission in psychotic disorders that co-occur with substance use. Schizophr Bull. 2006;32(4):618–625. doi:10.1093/schbul/sbl007

45. Botteron K, Carter C, Castellanos FX, et al. Consensus report of the APA work group on neuroimaging markers of psychiatric disorders. Am Psychiatr Assoc. Published online 2012. https://www.researchgate.net/profile/Karen-Seymour/publication/261507750_Consensus_Report_of_the_APA_Work_Group_on_Neuroimaging_Markers_of_Psychiatric_Disorders/links/0c9605346a4d865d9b000000/Consensus-Report-of-the-APA-Work-Group-on-Neuroimaging-Markers-of-Psychiatric-Disorders.pdf

46. SCORE2 and SCORE2-OP. Accessed December 16, 2022. https://www.escardio.org/Education/Practice-Tools/CVD-prevention-toolbox/SCORE-Risk-Charts

47. de Winter MA, Büller HR, Carrier M, et al. Recurrent venous thromboembolism and bleeding with extended anticoagulation: the VTE-PREDICT risk score. Eur Heart J. Published online January 17, 2023. doi:10.1093/eurheartj/ehac776

48. 48. Kia SM, Rad NM, van Opstal D, et al. PROMISSING: Pruning Missing Values in Neural Networks. arXiv *[csLG]*. Published online June 3, 2022. http://arxiv.org/abs/2206.01640

49. Baytas IM, Xiao C, Zhang X, Wang F, Jain AK, Zhou J. Patient Subtyping via Time-Aware LSTM Networks. In: Proceedings of the 23rd ACM SIGKDD International Conference on Knowledge Discovery and Data Mining. KDD’17. Association for Computing Machinery; 2017:65–74. doi:10.1145/3097983.3097997

50. Wilcoxon F. Individual Comparisons by Ranking Methods. In: Kotz S, Johnson NL, eds. Breakthroughs in Statistics: Methodology and Distribution. Springer New York; 1992:196–202. doi:10.1007/978-1-4612-4380-9_16

